# The epidemiology of antibiotic-resistant clinical pathogens in Uganda

**DOI:** 10.1101/2023.10.28.23297715

**Authors:** Ritah Namusoosa, Ibrahimm Mugerwa, Keneth Iceland Kasozi, Allan Muruta, Grace Najjuka, Winifred D. Atuhaire, Susan Nabadda, Henry Mwebesa, Olaro Charlse, Isaac Ssewanyana, Aloysious Ssemaganda, Adrian Muwonge

## Abstract

**Background:** Antibiotic resistance (ABR) is silent global pandemic. Our current global control strategies are informed by evidence primarily from surveillance strategies. Here, we use a national surveillance dataset to demonstrate how such evidence can be systematically generated, in so doing we characterize ABR profiles of priority clinical pathogens and identify potential drivers in addition to inferences on antibiotic usage in Uganda.

**Results:** of the 12,262 samples cultured between 2019-2021, 9,033 with complete metadata were analyzed, Female patients accounted for 57.8% of the patients who were between 1 to 104 years. The isolated bacteria (69%) were clustered into twelve clinical pathogens and eight syndromes. This included *Escherichia coli* 26%(n=1620), *Klebsiella spp*.11% (n=685), *Acinetobacter spp*. 4%(n=250), *Citrobacter spp*. 4% (n=250), *Pseudomonas spp*. 3% (n=187), and *Enterobacter spp*. 2% (n=124), as well as *Salmonella spp*. 1% (n=62). Among gram-positive bacteria, *Staphylococcus aureus* 10% (n=623), *Enterococcus spp*. 8% (n=498), and *Streptococcus spp*. 2% (n=124) were predominant. *Acinetobacter baumannii* was predominantly multi drug resistant (MDR) and mostly recovered from septic wound infection (SWI). *Pseudomonas aeruginosa*, *Escherichia coli*, *Klebsiella pneumoniae*, and *Staphylococcus aureus* were also linked to ABR SWIs & urinary tract infections (UTIs). Male patients were more likely to carry ABR pathogens OR=1.14, 95% CI [1.12-1.42], p<0.001), within specific age groups (51-60, OR=1.16, 95% CI [0.88-1.28], p=0.001). Seasonality also influences ABR associated to clinical syndromes, for example, the second quarter, OR=2.1, 95% CI [1.9-2.6], p<0.001), is associated with resistance to narrow spectrum antibiotics OR=1,64, 95% CI [1.39-1.94], p<0.001) targeting respiratory tract infections (RTIs). ABR associated bloodstream infections (BSIs) were significantly more common than UTIs and RTIs.

**Conclusions:** ABR across clinical pathogens was increasing at a rate of 2.8% per year, with an upsurge in 2021. SWIs account for the disproportionately high prevalence of ABR and MDR mostly caused by *Acinetobacter spp. Staphylococcus aureus* including MRSA is main driver of BSIs. Male patients are far more likely to carry ABR in their adult life. Encouragingly, carbapenem resistance remains relatively low in-line with the predicted antibiotic use. Such evidence is critical for effective implementation and evaluation AMR National action plans, therefore national public health institutes (NPHIs) ought to invest in building capacity for surveillance and data analysis to support informed decision-making.

## INTRODUCTION

Antibiotic resistance (ABR) refers to the phenomenon in which bacterial organisms become unresponsive to active pharmaceutical molecules intended for therapeutic purposes in hosts. The mechanisms underlying this process can be categorized into three types: a) intrinsic mechanisms, where a bacterial species is naturally resistant to a certain antibiotic or antibiotic family such as sporogenic *Bacillus cereus* (1), b) acquired mechanisms, which involve transfer through mobile genetic elements (2), and c) fitness-improving mutations in genes encoding resistance (3). Globally, ABR is estimated to cause 1.27 million deaths (4), with Sub-Saharan Africa experiencing an average of 27.3 deaths per 100,000 persons attributable to ABR, indicating a higher burden of disease in these regions (5). Vulnerable populations, including neonates, infants, and pregnant mothers, bear a disproportionate level of this burden (5–7). The emergence of ABR is strongly associated with the level of antibiotic exposure and worryingly, the use of antibiotics in both humans and animals has increased by 46% over the past two decades (8), with a projected 67% increase in animal antibiotic usage by 2030 due to rapid climate-driven environmental changes (9). This level of use will inevitably come with an increase in ABR and extra-burden to already struggling health care systems especially in the developing world, which is why robust antibiotic stewardship strategies are urgently required to preserve the effectiveness of currently available antibiotics and safeguard public health (10,11). Global efforts to address antimicrobial resistance (AMR) as a whole have been gaining momentum since the launch of an action plan in 2015, which is implemented through national action plans (12). Among the pillars of these plans, generating and sharing evidence on the trends and drivers of AMR is critical as it provides evidence that enables stakeholders not only to customize interventions but also to strategically allocate limited resources (12). Therefore, in addition to research valuable insights into ABR dynamics can be obtained from national passive and active surveillance systems. In countries like Uganda, surveillance efforts are structured to encompass various syndromes such as respiratory, bloodstream, gastrointestinal, and urogenital infections, thereby linking ABR to specific pathogens (13).

Respiratory tract infections (RTIs) are often characterized by prolonged clinical episodes and, in some cases, excessive antibiotic exposure (14). In Uganda, *Streptococcus pneumoniae* is one of the major causes of community-acquired respiratory tract infections, which are often highly resistant to trimethoprim-sulfamethoxazole (TMPS) (15). Prior to the coronavirus disease 2019 (COVID-19) pandemic, the prescribing rate of antibiotics for RTIs in Uganda was estimated to be as high as 77.6%, with an average of 2.47 drugs per encounter (16). Indeed, this high level of prescription is common in other parts of Africa and expected to have worsened during the pandemic (17–19). It is therefore plausible that this trend in antibiotic usage has influenced the prevalence of AMR associated with RTIs in these settings.

Urinary tract infections (UTIs), although associated with age and gender, females account for a significant proportion of bacterial recovery and antimicrobial resistance at various surveillance sentinel sites in Uganda (20,21). Here *Escherichia coli* and *Klebsiella pneumoniae* have been reported as the main culprits, linked to high resistance to second-line drugs such as ciprofloxacin and amoxicillin (22). The trends in UTIs UTI-associated resistance are also true for extended-spectrum beta-lactamase producers). Between 2014 and 2018, *Acinetobacter species* and *Staphylococcus aureus* were responsible for 17.5% and 12.4% of surgical wound infections (SWIs), respectively, at Mulago National Referral Hospital, with most of the cases associated with multiple drug resistance (24). A recent study predominantly focusing on sepsis among adults revealed that the prevailing sources of bloodstream infections (BSIs) are mainly due to *cytomegalovirus*, *Mycobacterium tuberculosis*, *Plasmodium*, and *Streptococcus pneumoniae* (25). In contrast, a survey of pediatric BSIs within rural settings highlighted that the predominant causative agents were *Klebsiella pneumoniae*, *E. coli*, and *Staphylococcus aureus* (26). These findings, along with other anecdotal evidence from literature, suggest that these syndromes represent the greatest burden of AMR in Uganda which the National Action Plans (NAPs) together with the associated surveillance plans for both drug-bug resistance and antimicrobial use and consumption aim to address. However, the primary challenge for the NAPs is the limited availability of the required resources to generate evidence for the guidance towards the strategic implementation and evaluation. Here, AMR surveillance active or passive surveillance strategies can serve as critical sources of evidence for NAPs implementation and outcome realization. In our earlier country situation report, we presented Uganda’s efforts towards developing a sentinel surveillance system (27) which is a non-trivial endeavor, especially in Low- and Middle-Income countries (LMIC) where data quality is affected by manual data capture methods, inadequate diagnostic capacity, inadequate resources including both financial and human resources (11).

Without quantifying the AMR burden, it remains difficult to control as NAPs cannot direct resources where they are critically needed. As such implementation and evaluation of such efforts becomes severely limited. Here we utilize the most comprehensive phenotypic, clinical and demographic AMR surveillance dataset from Uganda to examine the diagnostic characteristics and ABR of clinical pathogenic bacteria to identify the factors associated with ABR between 2019 and 2021 with the overarching goal of demonstrating the potential utility of sentinel surveillance data in informing and shaping the implementation of AMR national, regional and global action plans to combat AMR.

## 2.0 Materials and Methods

### 2.1 Study Design and Setting

This was a retrospective analytical study using National Microbiology Reference Laboratory data obtained from the sentinel site surveillance system for antibiotic sensitivity profiles of clinical pathogens in Uganda between 2019 and 2021. These data were collected through the national surveillance network of microbiology laboratories (**Figure S1a**), utilizing the electronic laboratory information management system from the African Laboratory Information System (ALIS) in Uganda (**Figure S1b**). No patient identifying variables are used for this process, so variables such as age, sex, district, health facility visited, and culture results are used in addition to the antimicrobial susceptibility test results for significant monomorphic bacterial organisms. Notably, for urine samples, a significant organism was identified as the growth of > 10^4^ CFU/ml of monomorphic organisms.

#### Sample size estimation

A total of 12,262 records were available in the ALIS database over the three-year period from 2019 to 2021. Of these, 9,033 records from fifteen sentinel sites were included in the analysis following data cleaning and elimination of incomplete entries, duplicated entries and entries without antibiotic susceptibility test results. (**Figure 1**).

**Figure 1.**
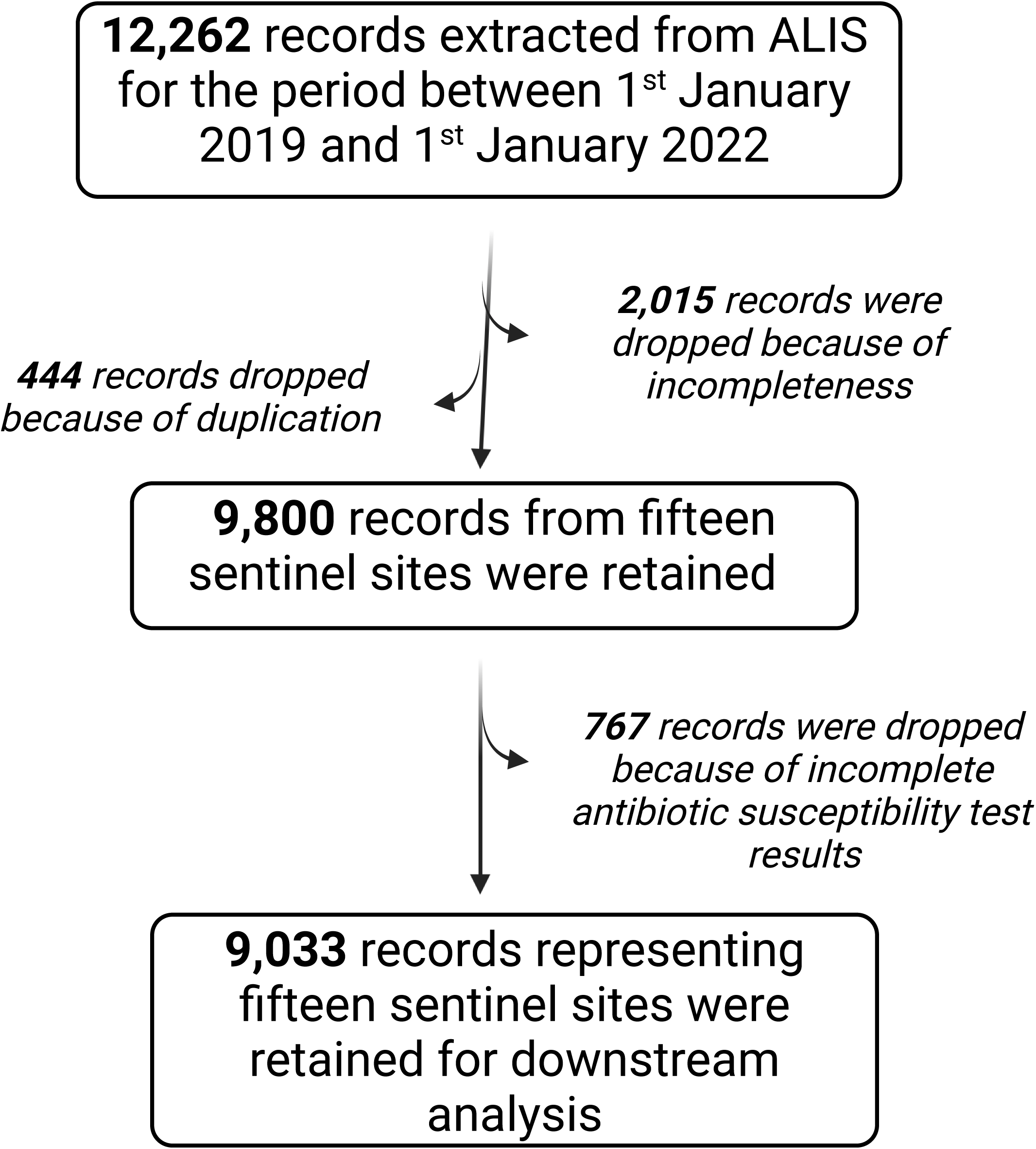
Screening protocol for datasets in the National Laboratory Information system.

### 2.2 Specimen collection

Specimen which included blood, urogenital swabs, respiratory samples, aspirates, cerebrospinal fluid, pus, and wound swabs were collected from both outpatient and inpatient departments at AMR sentinel sites under aseptic conditions (28) for microbiological culture to identify pathogens and for Antimicrobial Susceptibility Testing (AST).

### 2.3 Specimen processing and antimicrobial susceptibility testing

Aspirates, cerebrospinal fluid, urogenital and wound swabs, respiratory samples as well as pus swabs were initially inoculated on blood agar, chocolate agar, and MacConkey agar and samples showing significant growth were subsequently subjected to gram staining. Urine specimens collected in universal wide-mouthed sterile urine cups were inoculated on CLED (cysteine-lactose-electrolyte deficient) agar using calibrated loops. Samples with a significant growth of ≥ 104 CFU/ml on CLED agar were then subjected to gram staining. Following gram staining, Gram-negative colonies were subcultured on MacConkey agar, while gram-positive colonies were subcultured on blood agar.

Blood samples were aseptically collected in BD blood culture bottles (BD BACTEC™) and incubated in the BD BACTEC™ blood culture machines at the facilities. Those that turned positive were then gram stained and subcultured on blood agar, chocolate agar and MacConkey agar.

Following isolation, Gram-negative organisms were identified using tests such as triple sugar iron agar, urea, Simmons citrate agar, sulfur indole motility test, and oxidase test while gram-positive organisms were identified using tests like catalase, coagulase, DNase, bile esculin hydrolysis, optochin sensitivity test, and novobiocin sensitivity test (**Figure S2**).

Antimicrobial susceptibility tests (AST) were done on Mueller Hinton agar using the Kirby-Bauer disc diffusion method where sensitivity was obtained by measuring the zone diameters of clearance. Antibiotic discs were selected, and results interpreted following the CLSI (Clinical Laboratory Standard Institute) guidelines. The resulting identified culture isolates were then transported from the sentinel sites to the National Microbiology Reference Laboratory via the National Sample Transport network for cataloguing and retesting to confirm the reported phenotype and AST results.

All associated specimen metadata was recorded in microbiology laboratory register as well as the electronic laboratory information management system from the African Laboratory Information System (ALIS). Data variables captured included age, sex, district, health facility visited, culture results as well as antimicrobial susceptibility test results for significant monomorphic bacterial organisms. The laboratory generated data) was then merged with the metadata for the downstream analysis. No patient identifying variables were included in this analysis.

### 2.4 Statistical Analysis

The analysis was performed using an anonymized dataset exported from ALIS, containing metadata variables as shown in (**Table S1**). . Therefore, our initial investigation aimed to assess the impact of the COVID-19 pandemic on the national AMR surveillance system. To address this, we compared the trends in culture results with the cumulative burden of COVID-19 cases during the same timeframe. Furthermore, we conducted an analysis of the incidence of gram-negative and gram-positive pathogens reported per 100,000 population, categorized into quarterly intervals each year.

To visually represent changes in resistance, we plot distributions of raw disc diffusion diameters for twenty-five pathogens on thirty-one antibiotics over a span of three years. The interpretation of these plots is as follows: if the distribution shifts towards the left-over time, it suggests an increasing trend in resistance, whereas a shift towards the right suggests a trend of increasing susceptibility.

We then compared the prevalence of antibiotic resistant pathogens, including monoresistance and multi-resistance patterns, across variables such as gender, age, location, and clinical syndrome of the patients. This enabled us to identify clusters based on temporal, spatial, and patient-related factors. Our second and third research questions were to determine the factors associated with mono drug and multidrug resistance in Uganda. We run several mixed effects logistic models on data frames subsets by a) pathogens, i.e., key gram positive and negative pathogen, b) antibiotic classes and c) the clinical syndromes from which pathogens are recovered. This approach allowed us to appreciate the differential explanatory power of factors such as gender, age group’s location and the temporal effect. The models are developed and implemented using the lme4 package in R (Version 4.1.1).

The response variable in our model was the monoresistance status of the bacteria, indicating whether they were resistant or susceptible to a particular antibiotic, or the presence of multi-resistance. Multi-resistance was defined as resistance to at least three clinically relevant antibiotics for a given pathogen.

The explanatory variables used in the model are listed (**Table S2 & S3**), and the random variable was considered. Associations between the response variables and explanatory variables, as well as their interactions, were tested using likelihood ratio tests (LRTs) with the drop1 function in the lme4 package. The model selection process began with a complex model and employed backward stepwise variable selection, concluding with the simplest model based on the Akaike information criteria. Assumptions such as linearity, multicollinearity, and independence were assessed. Linearity was evaluated by examining residuals and fitted values of a variable, multicollinearity was checked using the variance inflation factor, and independence was assessed by comparing the correlation of group means to the effects. Normality of residuals was determined using Q-Q plots when applicable. Only variables with significant effects (P<0.05) were retained in the final model.

Finally, as none of the metadata factors included antibiotic use by patients, we relied on the established notional relationship between Antimicrobial Use (AMU) and Antimicrobial Resistance (AMR) to generate co-occurrence networks per pathogen and syndrome, serving as a proxy for antibiotic use. We constructed a binary matrix (1, 0) where “1” indicates resistance and “0” susceptibility to antibiotics respectively. Subsequently, a network was created using the igraph package (version 1.5.1) in R, where the node and edge thickness represent antibiotic and the frequency of connections between two antibiotics, respectively. The edge thickness is directly proportional to the frequency of their co-occurrence. Thus, the most frequently used antibiotics for a given pathogen can be inferred.

## 3.0 Results

### 3.1 Descriptive summary of study population

We analyzed the output of AMR surveillance conducted between 2019 and 2021. During this period, a total of 12,262 samples were submitted for culture across fifteen sentinel sites. 9033 samples were used for this analysis, representing (57.8%) females and an age range (1 to 104 years) (**Figure S3 and Figure 4a)**. From these samples, a total of (6284) bacteria and (738) Yeast was recovered, indicating an average culture recovery rate of (72.7%). The recovery rate steadily increased until the first quarter of 2021, here recovery of gram-negative bacteria (74.7%) was disproportionately higher than gram-positive bacteria (25.2%) (Fig 2a, Table S1). At regional level, the recovery rate was highest in the mid-western and western regions (80%), and lowest in the central and northern regions (35%) (**Figure 2b**). Comparing the incidence rates and cumulative incidence of selected bacteria and COVID-19 cases respectively, reveals an inverse relationship (**Figure 2 c & d**). Furthermore, because the trend is the same across all selected bacteria it shows an effective reduction in samples collected per quarter as the COVID-19 pandemic size grew. Yeast species including *Candida spp.* and *Cryptococcus spp.* accounted for 11% of the recovered organisms. Among gram-negative bacteria we observed *Escherichia coli* (26%), *Klebsiella spp.* (11%), *Acinetobacter spp.* (4%), *Citrobacter spp.* (4%), *Pseudomonas spp.* (3%), *Enterobacter spp.* (2%), and *Salmonella spp.* (1%). On the other hand, *Staphylococcus aureus* (10%), *Coagulase negative Staphylococcus* (13%), *Enterococcus spp.* (8%), and *Streptococcus spp.* (2%) accounted for the gram-positive bacteria (**Table S4**).

**Figure 2.**
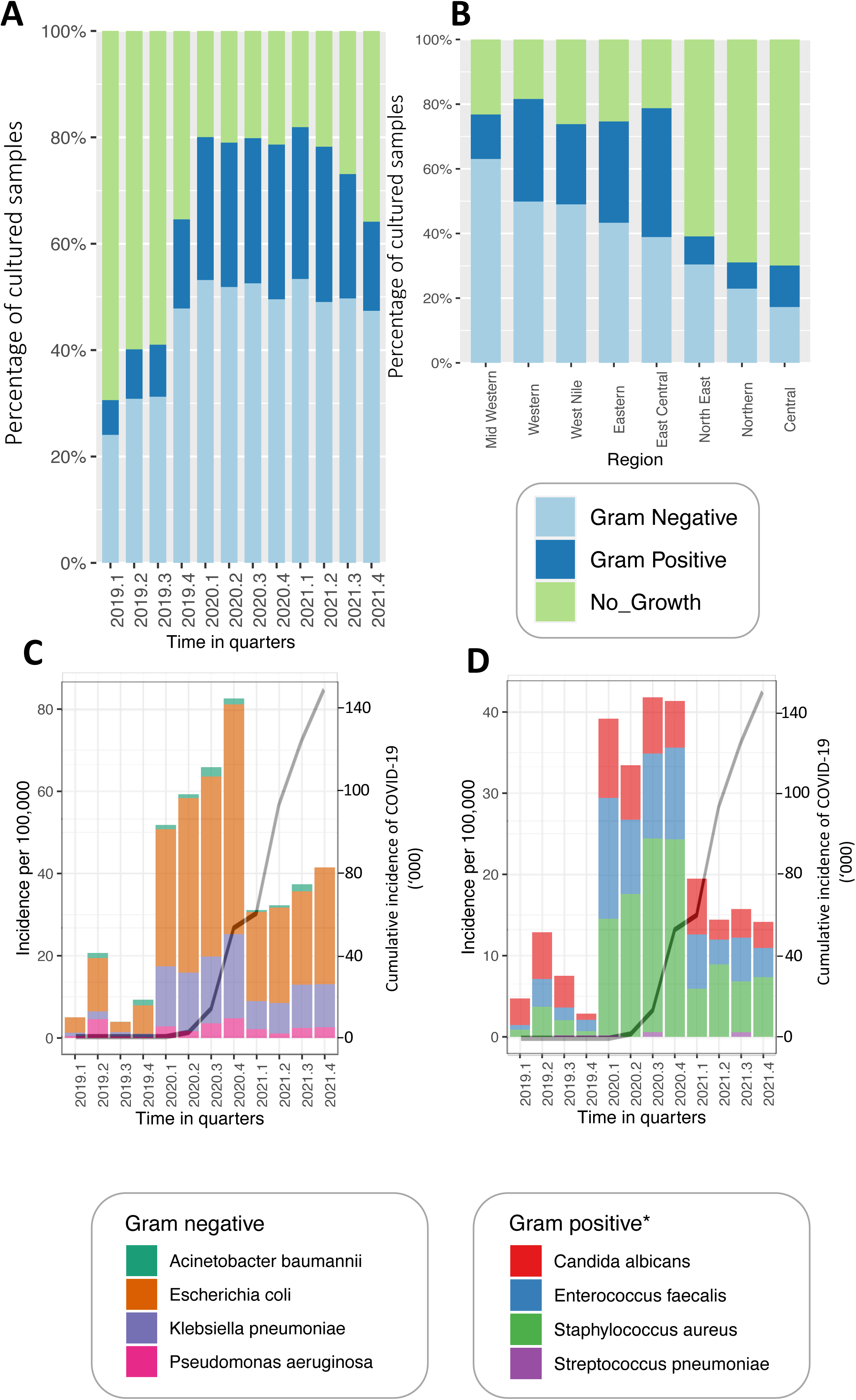
The characteristics and outputs of Uganda’s AMR national clinical surveillance system. Figure 1A & B, the culture recovery rate per quarter and region respectively. These show the proportion of bacteria recovery from the submitted samples between 2019-2021. Figure C&D show the incidence of gram negative and positive respectively. Note that we have included Candida within the comparison of the latter.

### 3.2 General trends in antibiotic resistance

By examining the distribution of disc diffusion diameters per antibiotic over the study period, we identified antibiotics that are overall gradually becoming less effective (**Figure 3**). This overall shift in the distribution is much more evident with watch category antibiotics such ceftazidime cefepime, meropenem, ertapenem and Vancomycin. In contrast, the trend observed for piperacillin-tazobactam indicates a reduction in resistance over time, given that the shift to the left was most in 2020 and then back to the right in 2021. We note a high level of resistance for beta-lactams, cephalosporins, including third and fourth-generation ceftriaxone and ceftazidime, as well as second-generation cefuroxime. A similar trend was observed for quinolones such as ciprofloxacin and levofloxacin.

**Figure 3.**
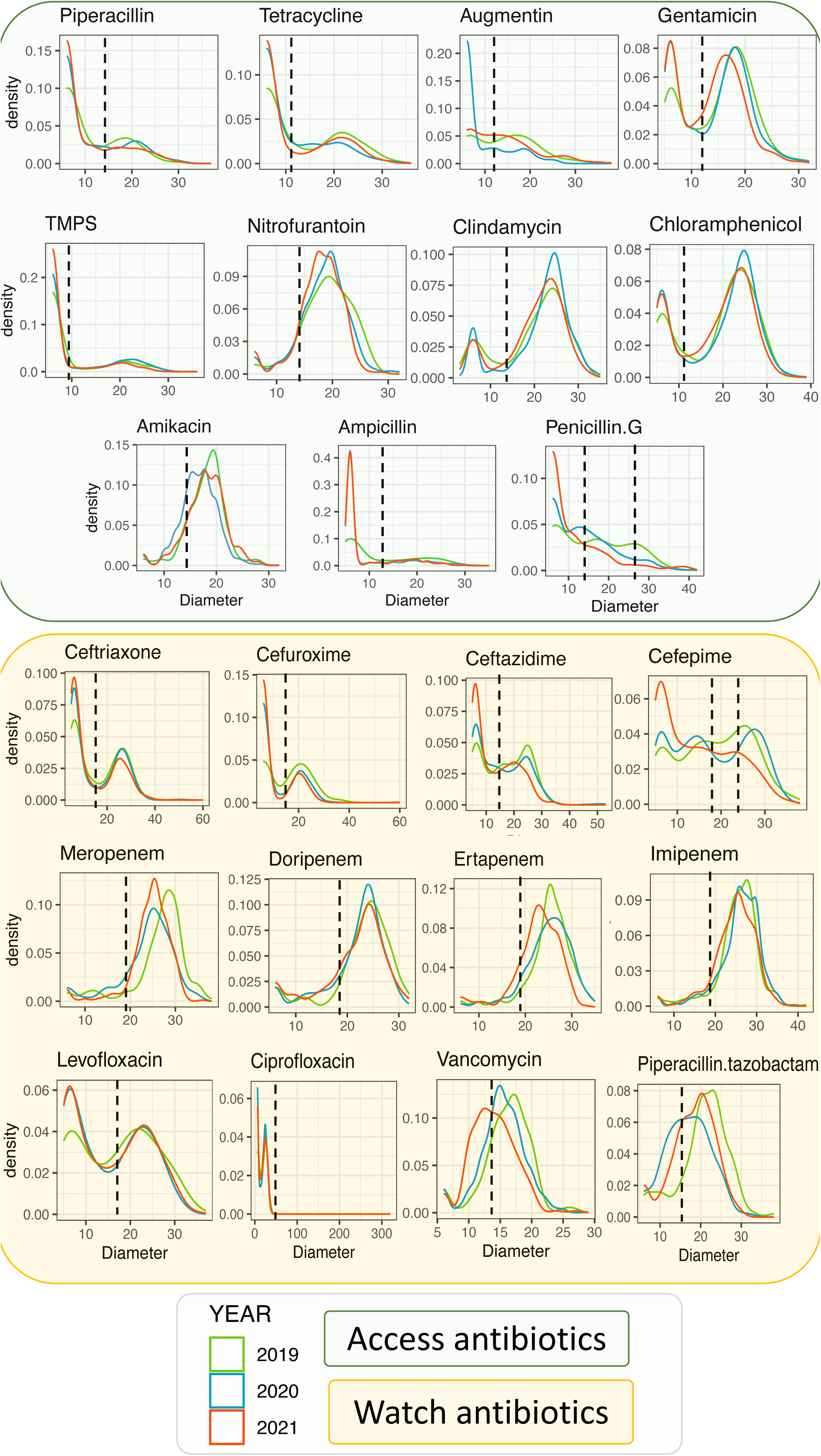
Shows the change in the distribution of disc diffusion diameter measurements over time for eleven and twelve access and reserve antibiotics respectively. The dotted line shows the threshold for resistance, revealing the extent to which ABR has increased during the study period

### 3.3 Patterns of antibiotic resistance against clinical pathogens

*Acinetobacter baumannii* were mostly resistant to more than one antibiotic (**Figure 4b**), and predominantly isolated from septic wound infection (SWI) samples. *P. aeruginosa*, *E. coli*, *K. pneumoniae*, and *S. aureus* that exhibited resistance to a single antibiotic were mainly recovered from SWI and UTI. We identify two populations of *E. coli* and *K. pneumoniae* represented by a bimodal distribution along the MDR gradient (**Figure 4b**). For example, *E. coli*, the group with extreme multi resistance was predominantly recovered from UTI and SWI samples. Across the study period, we noted a gradual increase in the prevalence of multidrug-resistant (MDR) and extensively drug-resistant (XDR) *A. baumannii, E. coli,* and *K. pneumoniae*. However, it is important to note that the prevalence of resistant *A. baumannii* (7%) was significantly lower compared to that of *E. coli* (29%) and *K. pneumoniae* (24%). On the other hand, we observe a considerably lower prevalence of MDR in *S. aureus* and *Enterococcus faecalis.* 10.7% of the former were MRSA, predominantly associated with SWIs in the western region of Uganda.

**Figure 4.**
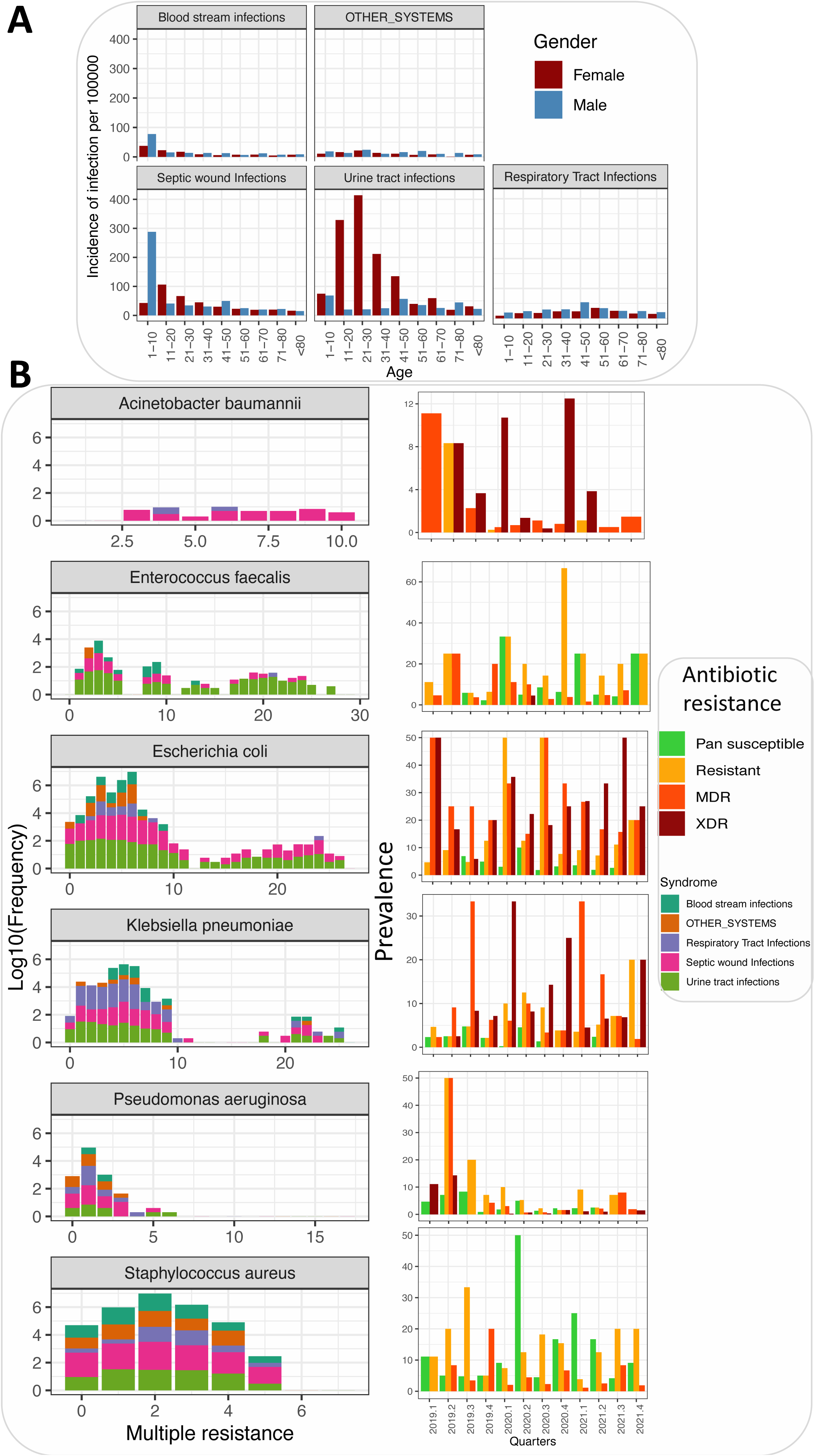
The incidence of clinical syndromes and their relationships with ABR. Figure 3A shows the incidence of clinical syndromes by age and gender, it is noteworthy that UTIs were more common among younger females. Figure 3B shows severity of ABR (x-axis the number of antibiotics to which the pathogen is resistant) depending on the syndrome. The second panel shows how this severity has been changing over time. For example, the prevalence of extremely resistant (>x antibiotics) *E. coli*, *Klebsiella Pneumoniae* and *Acinetobacter baumannii* has been increasing during this study period.

### 3.4 Factors associated with antibiotic resistance of clinical pathogens

We note some regional variation in ABR, for example, ABR was highest and lowest in the Northern (OR=2.26, 95%CI [2.1-2.8], P<0.001) and Northeast (OR=0.28, 95%CI [0.23-0.31], P<0.001) regions respectively. Comparatively, the levels of ABR were lower in the central region relative to eastern (OR=1.26, 95%CI [1.3-1.32], P < 0.001), mid-western (OR=1.56, 95%CI [1.1-1.61], P<0.001) and west Nile (OR=1.24, 95%CI [1.12-1.43], P=0.001).

#### 3.4.1 Age and gender associations with antibiotic resistance

The age and gender of patients were associated with ABR, however this varied depending on factors such as the syndrome, pathogen, and antibiotic class in question (**Figure 5**). Male patients were more likely to carry antibiotic resistant (ABR) pathogen than females especially if the pathogen in question was *Escherichia coli* (OR=1.18, 95%CI [1.12-1.43], P<0.001) or *Klebsiella pneumoniae* (OR=1.25, 95%CI [1.12-1.43], P<0.001) isolated from UTI samples (OR=1.36, 95%CI [1.12-1.43], P<0.001) and RTI (OR=1.53, 95%CI [1.12-1.43], P<0.001). This association was also evident if the antibiotic in question was a fluoroquinolone (OR=1.28, 95%CI [1.12-1.43], P<0.001) or cephalosporin (OR=1.19, 95%CI [1.12-1.43], P=0.006) (**Table S5** and S**6)**. Even when we consider a wide variety of explanatory variables this differential prevalence between gender remains statistically significant for male patients (OR=1.14, 95%CI [1.12-1.43], P<0.001). It is worth noting that a similar pattern is observed for Multi-Drug Resistance (MDR) (OR=1.47, 95%CI [1.12-1.43], P=0.007). In general, the prevalence of ABR tended to increase with age, with a significant increase observed among all age groups except for 11-20 and 41-50 (**Figure 6A**). Figure 5. Odd ratios on factors associated with syndromes, pathogens and antibiotics in the study area. A = Overall effect of the factors on resistance, B = Resistance across the major syndromes, C = Major bacterial and associated syndrome.

**Figure 5.**
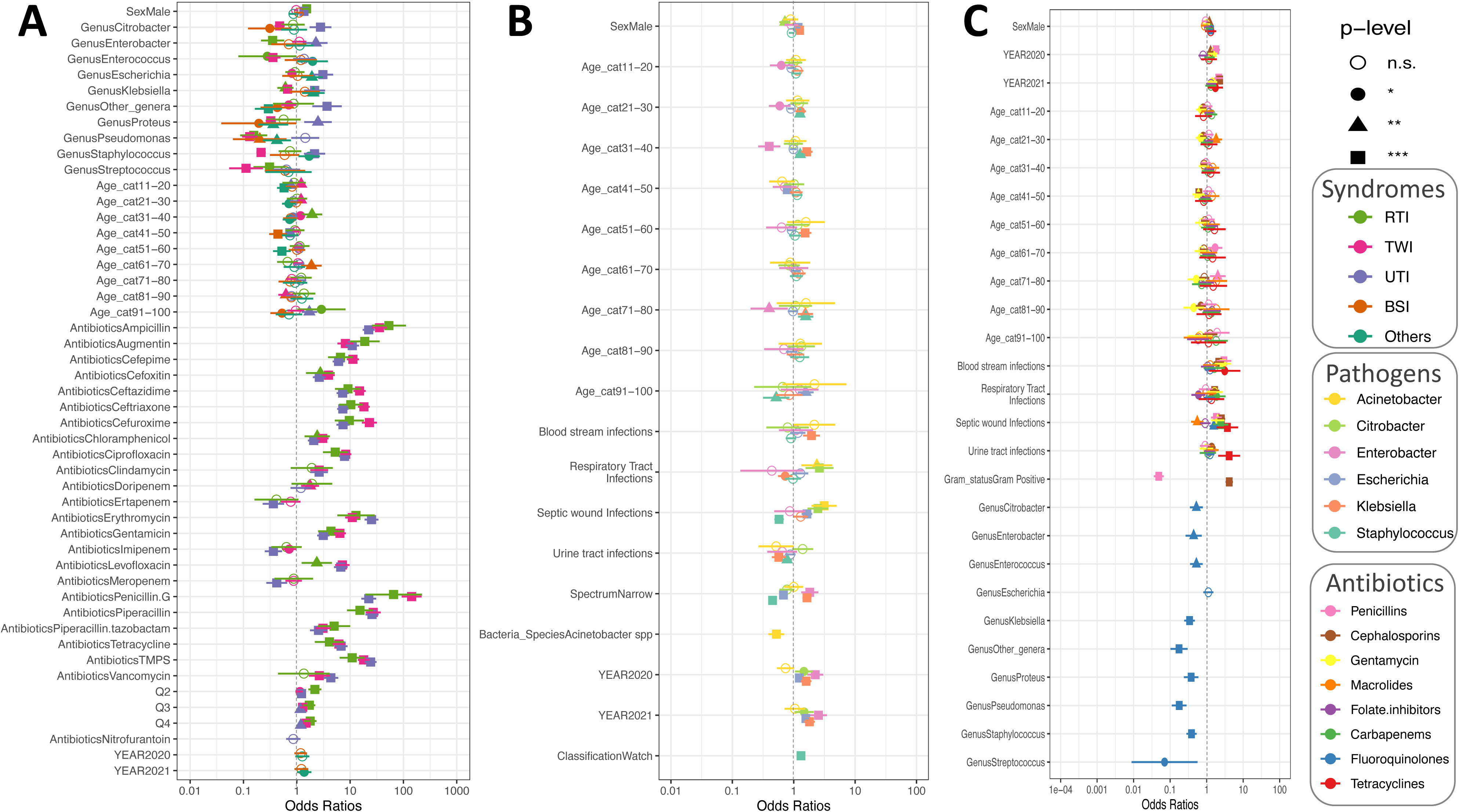
Using a total of 19 mixed effects logistic regression models (R2=10%-54%), we explore factors associated with ABR. Figure 4A compares five models of syndromes, Figure 4B compares six clinical pathogens and Figure 4C compares antibiotic classes. For all three comparisons male patients were more likely to carry ABR pathogens than females. The tabular output of these modes is presented in table S1-S6

**Figure 6:**
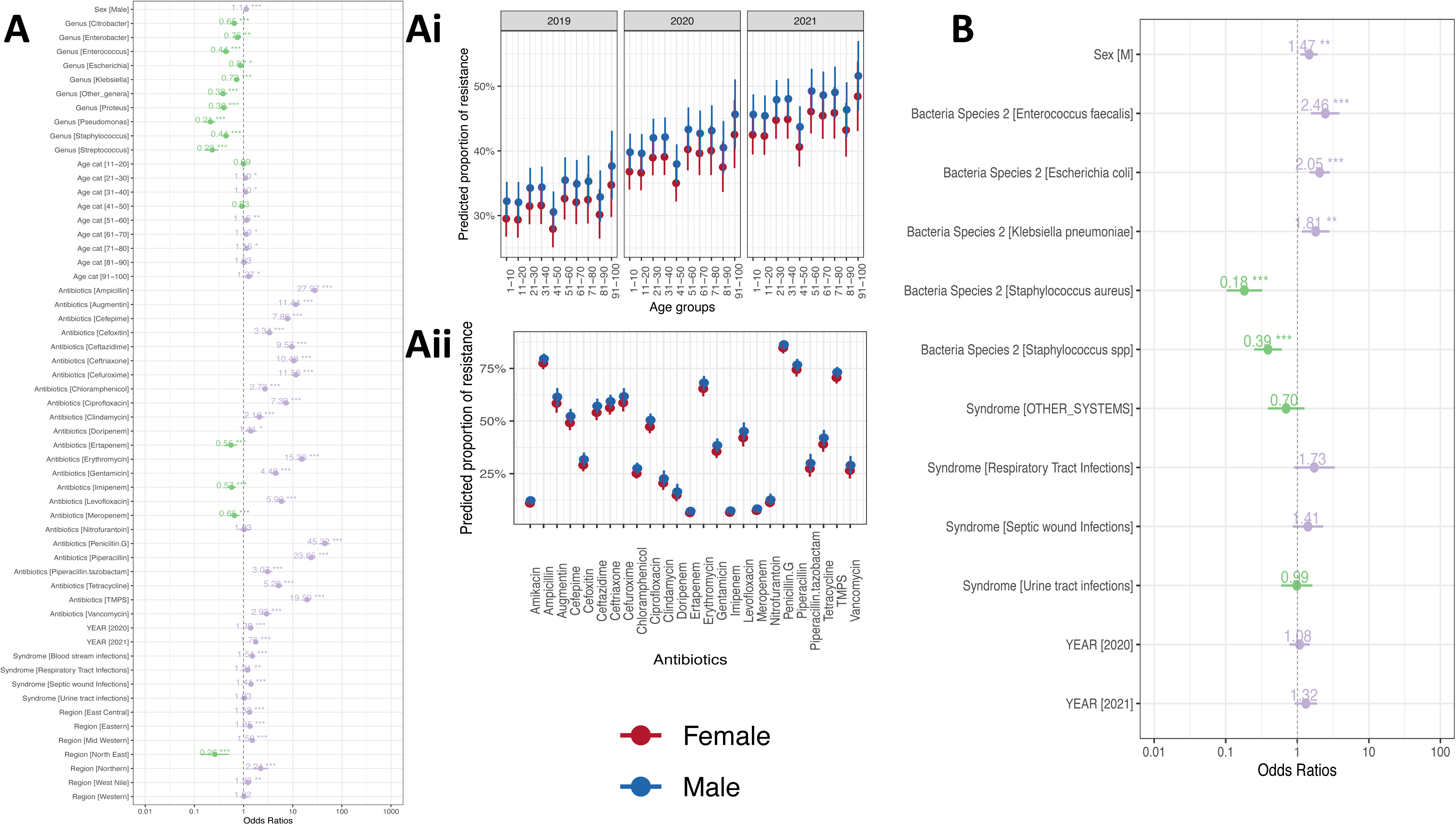
Full mixed effects logistic models examining the drivers of ABR. Figure A&B show the model output for mono and multidrug resistance. Figure Ai shows the steady increase of the model predicted ABR across age-groups and gender. Figure Aii is the predicted ABR per antibiotic.

#### 3.4.2 Clinical syndromes associated with antibiotic resistance

We observe that in general prevalence of ABR was lower and higher for pathogens recovered from the RTI and SWI for most antibiotics respectively. BSI-associated-ABR was less and more common among 41-50 (OR=0.44, 95%CI [0.30 – 0.65], P<0.001) and 61-70 (OR=1.88, 95%CI [1.21 – 2.92], P=0.005) age groups respectively when compared with 1–11-year-olds (**Table S7**). RTI associated ABR were more common among 31-40 (OR=1.93, 95%CI [1.24 – 3.00], P=0.003) age groups compared to 1–11-year-olds, and more likely to be males. Carbapenem resistant *Escherichia coli* and *Klebsiella pneumoniae* were more common with SWI (OR=2.4, 95%CI [1.23 – 4.56], P<0.001) and RTI (OR=1.86, 95%CI [1.12-1.43], P=0.034) (**Figure S5**). UTI associated ABR to Cephalosporins, Penicillin, Macrolides and Fluoroquinolones were more common among *Escherichia coli, Klebsiella pneumoniae, Proteus Mirabilis, Enterococcus faecium* than *Acinetobacter baumannii* (OR=0.49, 95%CI [1.12-1.43], P<0.001) (**Figure 5**). When we account for a full range of predictors, we note a higher prevalence of BSI associated ABR compared to UTI and RTIs (**Figure 6A**). Indeed, a similar relationship is shown between UTI and BSI for MDR prevalence (**Figure 6B**)

#### 3.4.3 Clinical pathogens and their associated antibiotic resistance profiles

ABR prevalence was disproportionately higher for *Acinetobacter baumannii,* but we also note significant variations across clinical pathogens. For example, RTI associated with ABR was less likely caused by *K. pneumoniae* (OR=0.61, 95%CI [0.43 – 0.87], P=0.006) and *Enterococcus faecalis* (OR=0.28, 95%CI [0.08 – 0.98] P=0.046) when compared to *Acinetobacter baumannii.* A similar relationship was noted for SWI but the opposite for UTIs, here ABR was more common for *Enterococcus faecalis* (OR=2.28, 95% CI [1.38 – 3.77] P<0.001)*, Escherichia coli* (OR=3.09, 95%CI [1.98 – 4.81], P<0.001) *and K. pneumoniae* (OR=2.16, 95%CI [1.37 – 3.39] P=0.001) (**Figure 5 A & B, Table S7**). Furthermore, *K. pneumoniae* recovered from BSIs, UTIs and RTIs were more likely to exhibit resistance than those recovered from SWI. Here the likelihood of resistance varied significantly depending on the antibiotic in question. For example, the difference in levels of resistance between SWIs and BSIs versus RTIs and UTIs was particularly prominent for beta-lactams and penicillin. *S. aureus* recovered from SWI were less likely to be resistant when compared to those from BSI. It is noteworthy that the observed associations remained statistically significant when we consider the full range of predictors (**Figure 6A**). *Acinetobacter baumannii* was more likely to express the MDR phenotype than *Staphylococcus aureus* (OR=0.18, P<0.001) but less likely than *Enterococcus faecalis* (OR=2.46, P<0.001), *Escherichia coli (*OR=2.05, 95%CI [1.26 – 3.33], P<0.001) and *K. pneumoniae* (OR=1.18, P=0.009) (**Figure 6 and Table S7**).

#### 3.4.4 Classes of antibiotics associated with resistance

The highest prevalence of ABR was with Penicillin such as Penicillin G and Ampicillin. Here the prevalence was higher among 61-70 (OR=1.70, 95%CI [1.08 – 2.67], P=0.023) and 71-80(OR=2.02, 95%CI [1.22 – 3.32], P=0.006) with BSI (OR=2.96,95%CI [1.85 – 4.72], P<0.001), and SWI (OR=1.84, 95%CI [1.31 – 2.58], P<0.001) when compared to other infections. The full model shows that overall resistance against penicillin, particularly Penicillin G, was significantly higher than most antibiotics tested (**Figure 6A** and **Figure S4** & **S5**). The resistance to Beta-lactams was higher among the female patients (OR=1.35, P=0.031) aged 51-60 (OR=1.65, 95% CI [1.00 – 2.71], P=0.049) in 2020 (OR=5.68, 95%CI [3.95 – 8.1] P<0.001) and 2021(OR=1.79, 95%CI [1.25 – 2.55], P=0.001).

Resistance to cephalosporins was significantly lower among patients aged 11-30 (OR=0.80, 95%CI [0.66 – 0.97], P=0.024), but more likely among male patients (OR=1.19, 95%CI [1.05 – 1.34] P=0.006) with BSI (OR=2.15, 95%CI [1.55 – 2.96], P<0.001). The levels of UTI related resistance to individual generations of cephalosporins, such as Cefepime (OR=6.09, 95% CI [4.71 – 7.87], P<0.001), Cefoxitin (OR=2.65, 95%CI [2.00 – 3.50], P<0.001), and Ceftriaxone (OR=7.30, 95%CI [5.72 – 9.30], P<0.001), were significantly higher than aminoglycosides such as amikacin (**Figure 6**). Resistance to macrolides, such as erythromycins was more common among patients aged 21-30 (OR=1.18,95%CI [1.24 – 2.63], P=0.002) and 71-80 (OR=2.02, 95%CI [1.22 – 3.32], P=0.006) with SWI (OR=1.55, 95%CI [0.37 – 0.81], P=0.003). Similarly, carbapenem resistance was associated with RTIs (OR=1.86,95%CI [1.05 – 3.30], P=0.034) On the other hand, fluoroquinolone resistance was not associated with UTIs, but SWIs (OR=1.56, 95% [1.19 – 2.05], P=0.001). Finally, resistance to tetracycline was more common among patients with BSI (OR=3.21, 95%CI [1.17 – 8.31], P=0.023), SWI (OR=3.68, 95%CI [1.87 – 7.25], P<0.001) and UTI (OR=4.11, 95%CI [2.06 – 8.20], P<0.001).

#### 3.4.5 Temporal characteristics of antibiotic resistance in Uganda

We have observed a consistent increase of ABR across various clinical pathogens against a range of antibiotics during the study period, with a surge in 2021 (OR=1.66, P<0.001) relative to 2019. In this period, we also observe that this increase has been more pronounced for some antibiotic classes such as cephalosporins (OR=2.27, 95% [1.88 – 2.75], P<0.001) and tetracycline (OR=1.73, 95% [1.07 – 2.79], P=0.024). Finally, we detected a seasonality signal associated with ABR, but this was a more pronounced feature for RTI, with consistently high levels of ABR in the second quarter of each year (OR=2.18, 95%CI [1.65 – 2.90], P<0.001), which indeed continues to increase across the rest of the quarters. To some extent the same was true for SWI, (OR=1.15, 95%CI [1.02 – 1.29], P<0.001), UTIs (OR=1.23, 95%CI [1.11 – 1.36], P<0.001)

### 3.5 Inferring antibiotic use patterns from ABR co-occurrence networks

The ABR co-occurrence patterns for *A. baumannii* suggest use of antibiotics is mainly to treat SWIs and the most used antibiotics here include ciprofloxacin cefepime, gentamicin and ceftazidime. The patterns for *E. coli* and *K. pneumoniae* show a difference in why antibiotics may be used, for the former use is mainly for managing SWIs and UTIs, while for the latter use is for managing a variety of syndromes as shown in (**Figure 7**). There is considerable overlap in the antibiotics mostly used to manage these two pathogens which include; ceftriaxone, ciprofloxacin, ampicillin TMPs, ceftazidime etc. The syndrome profile and likely antibiotic use for *Citrobacter freundii* is like that of *E. coli* (**Figure 7**). For gram positive pathogens such as *Staphylococcus aureus* the inferred antibiotic use maps to various syndromes, here antibiotic include penicillin G, erythromycin, ciprofloxacin and TMPS. The least used antibiotics to manage gram positive bacteria include quinolones such as levofloxacin and aminoglycosides such as Amikacin. Interestingly as a way of validating these inferences we note that vancomycin is comparatively the least used antibiotic and exclusive to gram positive.

**Figure 7:**
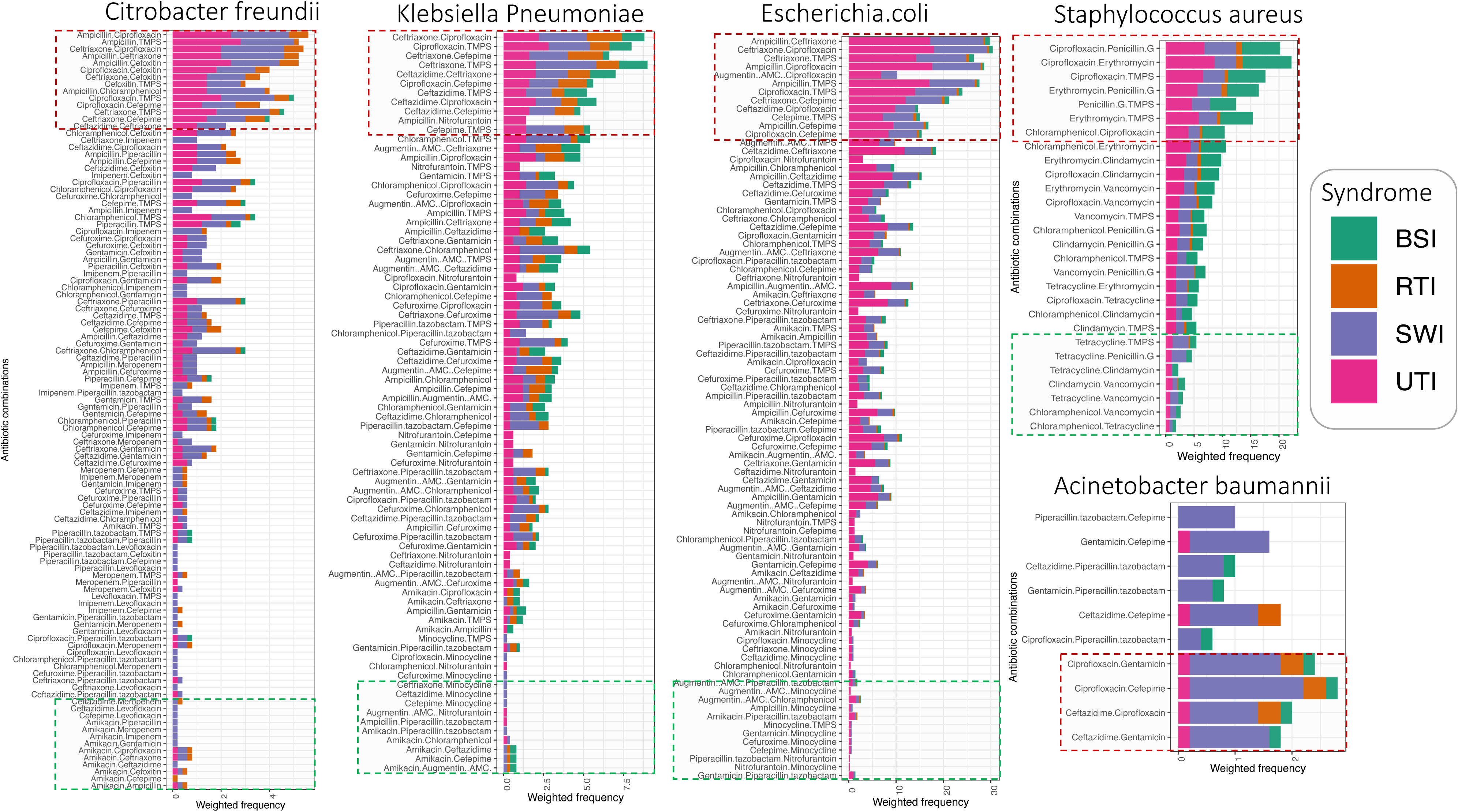
ABR co-occurrence patterns (CCP) are used here to infer antibiotic use patterns among selected clinical bacteria pathogens. The x-axis shows the CCP of the antibiotic and the y axis shows the weighted frequency. These are filled based on the syndrome, thereby showing the syndrome a given CCP is associated with. The dotted annotations show the topmost and least frequent pairs

**Figure 8:**
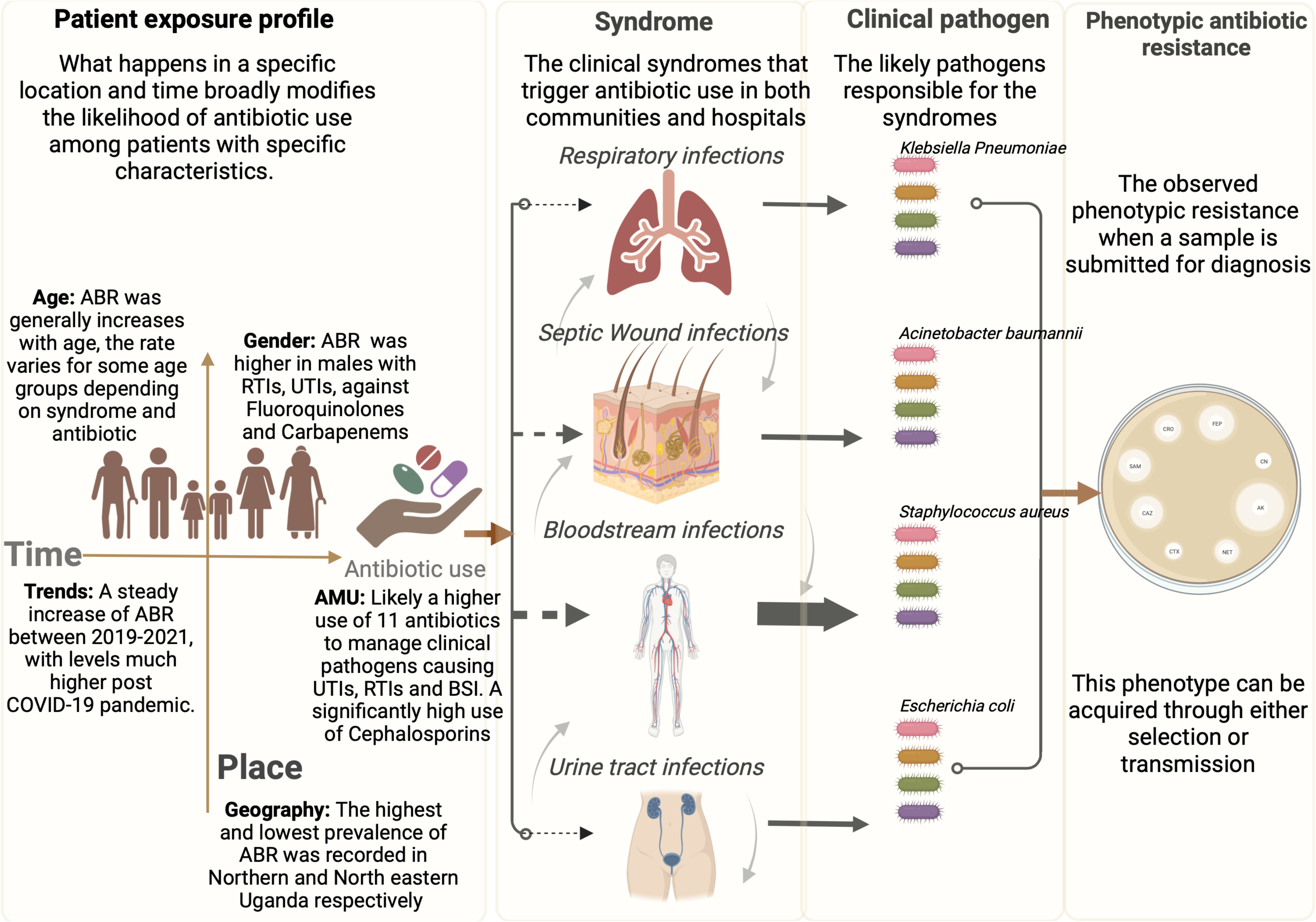
The inferred causal framework of ABR in Uganda based on the modeling output. In Uganda, *K. pneumoniae, A. baumannii, S. aureus*, and *E. coli* were major pathogens observed in the population.

## 4. DISCUSSION

In this study, we investigated the drivers of antibiotic resistance exhibited by common clinical pathogens in Uganda using the National AMR surveillance dataset between 2019 to 2021. The overarching goal was to underscore the potential utility of surveillance data in informing and shaping the implementation of global and national action plans to combat AMR (27). Our findings revealed a steady increase in the prevalence of antibiotic resistant (ABR) clinical pathogens between 2019 to 2021, with a notable surge in the last year. This finding also highlights the potential impact of COVID-19 on the quantitative and qualitative microbiological output of the national AMR surveillance system especially in the central region of Uganda (29). The findings also show how integral age and gender are to the prevalence of ABR, especially that which is linked to respiratory tract infections (RTI) and urinary tract infections (UTI) caused by *Klebsiella pneumoniae*, *Escherichia coli*, and *Citrobacter freundii*. Septic wound infections (SWIs) disproportionately account for a high level of ABR compared to UTIs, and BSI. Pathogens from such syndromes are predominantly resistant to Penicillin, such as Ampicillin and Penicillin G, while Carbapenems, such as ertapenem, imipenem, and doripenem, had relatively low resistance rates. As such these and more dynamics discussed below provide valuable insights into the complex and multi-level drivers of clinical antibiotic resistance and may serve as a foundation for informed decision-making during interventions and prioritization allocation for the national action plan.

### The impact of COVID-19 on the AMR sentinel surveillance system in Uganda

We observed both quantitative (number of samples processed) and qualitative (key performance indicators) reductions in the performance of the AMR National Sentinel Site Surveillance system at the Ministry of Health. This decline in performance coincided with the COVID-19 lockdown in the second quarter of 2020, indicating a plausible connection to increased demands on the healthcare system and a likely reallocation of funding and human resources. The decrease in key performance indicators, such as culture recovery rate, suggests challenges related to limited human resources and/or insufficient consumables. This situation is consistent with healthcare system challenges experienced elsewhere, both in developed countries (30) like Ireland and in developing countries (31) such as Ethiopia. These findings underscore the urgent need to bolster healthcare human resources, especially now when countries are formulating and testing future preparedness strategies.

Interestingly, the decline in both qualitative and quantitative performance did not correspond to a decrease in ABR levels. On the contrary, there was an observed increase in ABR levels, notably in 2021 compared to pre-pandemic times. This rise is likely attributable to the heightened use of antibiotics for treating and prophylaxis linked to COVID-19 (18,36). Essentially, this indicates a delay between usage and the observed impact on ABR. Assuming widespread use for COVID-19 began in the first quarter of 2020, we can estimate a lag of up to two quarters. When examined at quarterly level, it became evident that the surge in ABR associated with Respiratory Tract Infections (RTIs), Surgical Wound Infections (SWIs), and Urinary Tract Infections (UTIs) commences in the second quarter of each year and continues to rise consistently be that a different rates per syndrome. This pattern suggests a seasonal trigger, with the second quarter in Uganda being linked to seasonal influenza infections (15,38) and the onset of the rainy season (39). Hence, the ABR surge observed in 2021 could be attributed to both intrinsic seasonal factors and extrinsic selection pressures driven by imprudent antibiotic use associated with COVID-19. Alarmingly, these changes have led to a compounded average annual ABR increase of 2.8% across clinical pathogens and antibiotic classes. If not effectively addressed in Uganda, this trend could severely compromise the efficacy of common antibiotics by the year 2053.

### The role of age and gender on ABR prevalence

While the existing literature on gender-based differences in ABR remains limited, some reports suggest that ABR is more likely to be prevalent in females. This increased likelihood is attributed to the fact that females receive antibiotic prescriptions 27% more often in their lifetime compared to men (32). The perceived higher ABR risk in females stems from historical perceptions tied to biological factors, gender roles, and limited healthcare access. Paradoxically, these perceptions may introduce biases in how prescriptions are generated for females (40). In this study, the distribution of ABR among patients by gender appears to align with the notion of gender-based AMR risk but in opposition to the common knowledge. Here the risk appears to be skewed towards male patients, particularly when considering ABR for pathogens like *Klebsiella Pneumoniae* and *Escherichia coli* recovered from Respiratory Tract Infections (RTIs) and Urinary Tract Infections (UTIs). Notably, ABR is frequently observed against commonly used antibiotic classes such as fluoroquinolones (e.g., ciprofloxacin and levofloxacin) and cephalosporins (e.g., cefoxitin, ceftriaxone, and cefepime). Alarmingly, these male patients are also more likely to carry multidrug-resistant (MDR) pathogens. The reasons for this departure from existing literature are currently unknown, warranting a thorough investigation using longitudinal health and demographic surveillance survey resources. Considering health-seeking behavior, this divergence might imply that male patients tend to self-medicate within communities and only resort to visiting healthcare facilities when this approach proves ineffective. In this regard, recent studies on lower respiratory infections have concluded that male patients are 1.31 times more likely to succumb compared to females (16), while in India, male patients typically present with debilitating conditions such as diabetes and kidney stones, often associated with antibiotic overuse and abuse (41,42).

Furthermore, our findings agree with the widely accepted notion that lifetime exposure to antibiotics directly correlates with ABR (43), a link extensively documented in food animal production (44). Here we noticed a steady increase in ABR across age groups, although the relationship varied based on the syndrome, pathogen, and antibiotic in question. For instance, a disproportionately high occurrence of ABR *Klebsiella pneumoniae* was noted among 31–40-year-olds with RTIs. Conversely, a higher incidence of Cephalosporin, Penicillin G, and Vancomycin-resistant *K. pneumoniae* and *S. aureus* was observed among 11–30-year-olds with Surgical Wound Infections (SWIs) (45).

Tetracycline resistance is often regarded as legacy resistance due to its extensive use over the last 50 years (46). Our findings revealed significantly elevated levels of resistance among patients aged 51–60 years, supporting the concept of lifetime exposure.

In North America and Europe (47,48, 49), individuals aged 60 and above are 50% more likely to experience BSI (50,51). On the other hand, in Asian and African, BSI is more prevalent in pediatrics but prevalent across all age groups (52,53). Although our findings do not precisely quantify the extent of BSI overall, they do indicate that Antibiotic-Resistant BSI was higher among infants and patients aged 61–70. This likely emphasizes the extensive use of antibiotics in managing this life-threatening syndrome. Additionally, it’s plausible that this situation is exacerbated by compromised immune states associated with both age groups, often following HIV and TB infections (53,54). In reality, BSI poses a global challenge, affecting over half a million children, further exacerbated by the rise in Antimicrobial Resistance (55). In Uganda the disproportionate prevalence of AMR among BSI cases is of clinical concern and ought to have targeted interventions as part of the AMR national action plan.

### Syndromes associated with ABR among priority clinical pathogens

The global priority pathogen list by the World Health Organization has twelve bacteria strongly linked to major infections and high levels of ABR (56). In this study, we focus on eight out of the twelve bacteria and assess the ABR levels linked to various clinical syndromes. For example, UTIs and RTIs collectively account for a global burden of 150 million (57) and 17.2 million cases per year (58) respectively. Our findings reveal a growing prevalence of ABR associated with *E. coli* and *K. pneumoniae* recovered from these syndromes (59). While UTIs and RTIs were the most prevalent, BSI, especially those associated *Staphylococcus aureus*, *E. coli*, and *K. pneumoniae*, show disproportionately higher levels of ABR (60). In Europe, *Staphylococcus aureus* BSI is estimated at 20-30 per 100,000 population (61,62), surprisingly higher than previously reported levels in Malawi (63). This type of BSI is linked to serious prognoses, but the additional economic burden remains under investigation, particularly in low- and middle-income countries (LMICs) (64). In Uganda, the growing levels of Vancomycin-resistant *Staphylococcus aureus* and *Enterococcus faecalis* is likely to pose a significant risk to pediatric BSI (26) and surgical outcomes.

On a global scale, the incidence of BSI caused by *E. coli* and *K. pneumoniae* surpasses that of *S. aureus*, a trend which tends to be amplified by region-specific hypervirulent strains (61). Here we observed significantly high of ABR in *K. pneumoniae* and *E. coli*, particularly against narrow-spectrum antibiotics in the years 2020 and 2021. The international surveillance of these two pathogens primarily monitors resistance to fluoroquinolone, third-generation cephalosporin, and carbapenem. Our findings indicate substantial resistance levels against fluoroquinolones and cephalosporins but comparatively lower resistance against carbapenems. Hence, it is imperative for National Antimicrobial Programs (NAPs) to bolster efforts promoting the judicious use of carbapenems to sustain these low resistance levels and counter the adverse trends observed in the other two antibiotic classes. As a note to researchers, we need to articulate the practical value of this evidence to healthcare professionals who play a central role in generating the surveillance data that underpins it. This will not only build trust in the processes of data sharing but also lay the foundation on which antimicrobial stewardship initiatives developed in our countries.

Lastly, we noted high levels of ABR and MDR associated with septic wound infections (SWI) attributed to *Acinetobacter baumannii*. Despite limited reporting on this pathogen in Uganda, available data suggest it accounts for approximately 17.5% of SWI cases at the National Referral Hospital (24). Such cases are considered challenging to treat due to their multidrug resistance phenotype, including 3% to 35% Carbapenem-resistant *Acinetobacter baumannii* (CRAB) (65).

The effectiveness of *A. baumannii* as an opportunistic pathogen capable of causing Bloodstream Infections (BSIs), Urinary Tract Infections (UTIs), and SWIs is extensively documented in the literature (66,67). Therefore, its prominence in this study can likely be attributed to its diverse mechanisms of antibiotic resistance, including β-lactamase acquisition, aminoglycoside modification, permeability defects, upregulation of multidrug efflux pumps, and alterations in target sites (68).

### Inferences on antibiotic use in Uganda

There is a well-established consensus in the literature linking AMU directly to the development ABR (43,69). Hence, co-occurrence networks of antibiotic resistance can provide valuable insights into AMU. Building on this understanding, we can reasonably infer those eleven antibiotic molecules, namely ceftriaxone, cefoxitin, cefepime, ceftazidime, ciprofloxacin, ampicillin, and Trimethoprim-sulfamethoxazole (TMPs), that are likely the most used in the management of *E. coli*, *K. pneumoniae*, *A. baumannii*, and *Citrobacter freundii*. It is worth noting that in the case of *A. baumannii*, these antibiotic agents would primarily and almost exclusively be used for managing surgical wound infections (SWIs). On the other hand, infections caused by gram-positive pathogens like *Staphylococcus aureus* are more likely to be treated using Penicillin G, Erythromycin, Ciprofloxacin, Chloramphenicol, Trimethoprim-sulfamethoxazole (TMPs), and Vancomycin. Remarkably, our inferences closely align with the treatment guidelines outlined in the Uganda clinical guidelines of 2016 (70). Specifically, Trimethoprim-sulfamethoxazole (TMPs) which is recommended as the first-line treatment for uncomplicated syndromes, while Penicillin G as the established primary therapeutic choice for severe RTIs and BSI. In cases of severe BSI, UTIs, and RTIs, ceftriaxone is the preferred first-line medication and cefixime when complications arise. The UCG also recommends ciprofloxacin as a first-line drug for RTIs and a secondary option for treating uncomplicated UTIs.

### Limitations of this study

Some districts under their respective regional referral hospital laboratories had limited data which would have affected representativeness, this is because the National sentinel surveillance system for AMR was activated in 2018, therefore adoption across districts varied. We have overcome this problem by clustering the district into regions which allowed sufficient degrees of freedom for the analysis. Most of the isolates analyzed from the biorepository were collected from government facilities with less or few from the private health sector. Authors believe the study offers a basis to promote prospective GLASS activities in other regional centers and laboratories in the country and genomics for surveillance of AMR at national level to guide policy.

## CONCLUSION

Our analysis indicates an average annual increase of 2.8% in antibiotic resistance (ABR) across various clinical pathogens. Notably, there was a significant upsurge in 2021, likely attributed to excessive antibiotic usage during the COVID-19 pandemic. Furthermore, we identified a notable rise in ABR associated with respiratory tract infections (RTIs), possibly linked to the rainy season occurring in the second quarter of each year.

It is concerning that septic wound infections (SWIs) were primarily caused by *Acinetobacter spp*., which exhibited disproportionately high levels of ABR and multidrug resistance (MDR). Life-threatening bloodstream infections were predominantly due to ABR *Staphylococcus aureus*, but MRSA was linked to SWIs mostly reported from the western region. Among *K. pneumoniae* and *E. coli* isolated from RTIs and urinary tract infections (UTIs), fluoroquinolone and cephalosporin resistance were significantly high, particularly among male and specific patient age groups.

Encouragingly, resistance against carbapenems, critical for managing the majority of gram-negative clinical pathogens, had a relatively low prevalence. These findings underscore opportunities for targeted interventions for the AMR national action plan, and the potential utility of surveillance data in informing local regional and global efforts. Critically, our findings highlight the scale of the public health challenge Uganda faces in addition to existing health care challenges.

## DECLARATIONS

### Ethical considerations

This publication displays AMR public health surveillance work. The National AMR Coordination Center of the Department of National Health Laboratories and Diagnostic Services for the Ministry of Health is mandated to release these surveillance reports. The samples and isolates were obtained from the department’s public health and surveillance activities. The results of this publication are intended for public health and improving patient care and management with bacterial infections. All isolates analyzed were anonymized to their samples and original sentinel site. The study was protocol and ethics approved was obtained from the Uganda National Health Laboratory Services Research and Ethics Committee under (Reference Number UNHL-2023-68).

### Author contributions

Conceptualization and design of the study, R.N., I.M, A.M.; data collection, R.N.; formal data analysis and preparation of figures and tables, A.M.; drafting of manuscript, A.M., K.I.K., I.M., R.N.; writing, reviewing and editing, A.M., K.I.K., I.M., R.N., M.A., G.N., W.D.A., S.N., H.M., O.C., A.S., and I.S. All the authors have read and approved the final manuscript.

### Source of funding

Supporting the establishment of AMR Sentinel surveillance has involved multi-stake holders’ engagements and support right from establishing National AMR Governance structures at both national and subnational level. The Government of Uganda through its Ministry of Health department of Laboratories and Pharmacy have provided funds for both human resource, infrastructure and laboratory supplies. Furthermore, health development partners like Fleming Fund, Centers for Disease Control-CDC under the Global Health Security Agenda and other health implementing partners. AM is core funded at the Roslin Institute as part of his Chancellor’s fellowship

### Conflict of interest

The data analyzed and its content are part of the mandate and outcome role for the National AMR Coordination Center under the Department of Laboratories. Therefore, there is no conflict of interest to declare. This as well applies to other collaborating institutions with the authors of this publication.

### Availability of data and materials used in the study

The AMR sentinel surveillance is premised on the Ministry of Health National Microbiology Reference Laboratory (NMRL) and its network of National and Regional Referral Hospital Laboratories and University Laboratories (n = 21) all at slightly different levels of performance. All these laboratories, including some from private, are required to share isolates and quarterly microbiology data to the AMR-NCC for validation. These isolates are retested and routinely archived in the National Biorepository. The tools used to capture patient level data, demographics, laboratory and clinical findings is the National Microbiology Laboratory Request Form and its Results form. Further to this, the National microbiology laboratory has developed a Laboratory Information Management tool that has been customized to both national and sub-national microbiology needs. The data can be availed upon request, the application for data access will be reviewed by the relevant panel within the Ministry of Health of Uganda.

## Supplementary files

### List of Supplementary Figures

Figure S1a. Uganda sentinel site AMR surveillance system

Figure S1b. Cascade of the local health care system and how it is communicated with global stakeholders such as the World health organization.

Figure S2. Flow chart for bacteria identification.

Figure S3. The distribution of sentinel surveillance participants by age and gender

Figure S4: Predicted prevalence of AMR, visualized by age and predicted resistance colored by gender and faceted by clinical syndromes.

Figure S5: Predicted prevalence of AMR, visualized by age and predicted resistance colored by gender and faceted by antibiotic.

### List of Supplementary Tables

Table S1. The descriptive summary of the AMR sentinel surveillance of Uganda characterizing the culture recovery rate as a key diagnostics performance indicator.

Table S2. The output from mixed effects logistic regression model for gram positives Table S3. The output from mixed effects logistic regression model for gram negatives Table S4. Summary of culture recovery rates over 3 years

Table S5. Mixed effects logistic regression model where the outcome was status of phenotypic resistance (resistance or not resistant) and four major antibiotic classes

Table S6. Mixed effects logistic regression model where the outcome was status of phenotypic resistance and other antibiotic classes

Table S7. Mixed effects logistic regression model where the outcome was status of phenotypic resistance (resistance or not resistant)

## Data Availability

All data produced in the present study are available upon reasonable request to the authors

## Acknowledgment

We acknowledge the support of in-country implementing partners from the Infectious Diseases Institute and Baylor Uganda with support from the Fleming Fund and Centres for Disease Control. The National Microbiology Reference Laboratory and the National Coordination Center in implementing the National AMR surveillance program.

## Notes

### Competing Interest Statement

The authors have declared no competing interest.

### Funding Statement

This study did not receive any funding

### Author Declarations

The data used for this analysis is held by the Ministry of Health in Uganda, at the Uganda National Health Laboratories, which is mandated with AMR surveillance in the country. The data can be accessed, to do so one would need to make a request, which will be reviewed by a committee. A request can be made via the First Authors email or the corresponding email

## REFERENCES

1. Cox G, Wright GD. Intrinsic antibiotic resistance: Mechanisms, origins, challenges and solutions. Int J Med Microbiol [Internet]. 2013 Aug;303(6–7):287–92. Available from: https://linkinghub.elsevier.com/retrieve/pii/S1438422113000246

2. Lin J, Nishino K, Roberts MC, Tolmasky M, Aminov RI, Zhang L. Mechanisms of antibiotic resistance. Front Microbiol [Internet]. 2015 Feb 5;6. Available from: http://journal.frontiersin.org/Article/10.3389/fmicb.2015.00034/abstract

3. Marciano DC, Wang C, Hsu T-K, Bourquard T, Atri B, Nehring RB, et al. Evolutionary action of mutations reveals antimicrobial resistance genes in Escherichia coli. Nat Commun [Internet]. 2022 Jun 9;13(1):3189. Available from: https://www.nature.com/articles/s41467-022-30889-1

4. Murray CJ, Ikuta KS, Sharara F, Swetschinski L, Robles Aguilar G, Gray A, et al. Global burden of bacterial antimicrobial resistance in 2019: a systematic analysis. Lancet. 2022;

5. Taylor J, Hafner M, Yerushalmi E, Smith R, Bellasio J, Vardavas R, et al. Estimating the economic costs of antimicrobial resistance: Model and Results. RAND Corp [Internet]. 2014 Dec 10 [cited 2023 Jun 20];113. Available from: https://www.rand.org/pubs/research_reports/RR911.html

6. Mayanja R, Muwonge A, Aruhomukama D, Katabazi FA, Bbuye M, Kigozi E, et al. Source-tracking ESBL-producing bacteria at the maternity ward of Mulago hospital, Uganda. Aworh MK, editor. PLoS One [Internet]. 2023 Jun 8;18(6):e0286955. Available from: https://dx.plos.org/10.1371/journal.pone.0286955

7. Littmann J, Zorzet A, Cars O. Antimicrobial Resistance - A Threat to the World’s Sustainable Development - Dag Hammarskjöld Foundation. Ups J Med Sci [Internet]. 2016;121(3):159–64. Available from: http://www.daghammarskjold.se/publication/antimicrobial-resistance-threat-worlds-sustainable-development/

8. Browne AJ, Chipeta MG, Haines-Woodhouse G, Kumaran EPA, Hamadani BHK, Zaraa S, et al. Global antibiotic consumption and usage in humans, 2000–18: a spatial modelling study. Lancet Planet Heal [Internet]. 2021 Dec;5(12):e893–904. Available from: https://linkinghub.elsevier.com/retrieve/pii/S2542519621002801

9. Larsson DGJ, Flach CF. Antibiotic resistance in the environment. Nat Rev Microbiol. 2022;20(5):257–69.

10. Samreen, Ahmad I, Malak HA, Abulreesh HH. Environmental antimicrobial resistance and its drivers: a potential threat to public health. J Glob Antimicrob Resist [Internet]. 2021 Dec;27:101–11. Available from: https://linkinghub.elsevier.com/retrieve/pii/S2213716521001910

11. Kiggundu R, Lusaya E, Seni J, Waswa JP, Kakooza F, Tjipura D, et al. Identifying and addressing challenges to antimicrobial use surveillance in the human health sector in low- and middle-income countries: experiences and lessons learned from Tanzania and Uganda. Antimicrob Resist Infect Control [Internet]. 2023 Feb 9;12(1):9. Available from: https://aricjournal.biomedcentral.com/articles/10.1186/s13756-023-01213-3

12. WHO. Global action plan on antimicrobial resistance. World Heal Organ [Internet]. 2015;1–28. Available from: https://www.who.int/publications/i/item/9789241509763

13. UNAS, CDDEP G-U, Mpairwe Y, Wamala S. Antibiotic Resistance in Uganda : Situation Analysis and Recommendations [Internet]. Kampala, Uganda: Uganda National Academy of Sciences; Center for Disease Dynamics, Economics & Policy; 2015. 107 p. Available from: https://onehealthtrust.org/wp-content/uploads/2017/06/uganda_antibiotic_resistance_situation_reportgarp_uganda_0-1.pdf

14. Derbyshire EJ, Calder PC. Bronchiectasis—Could Immunonutrition Have a Role to Play in Future Management? Front Nutr [Internet]. 2021 Apr 29;8:652410. Available from: https://www.frontiersin.org/articles/10.3389/fnut.2021.652410/full

15. Rutebemberwa E, Mpeka B, Pariyo G, Peterson S, Mworozi E, Bwanga F, et al. High prevalence of antibiotic resistance in nasopharyngeal bacterial isolates from healthy children in rural Uganda: A cross-sectional study. Ups J Med Sci [Internet]. 2015 Oct 2;120(4):249–56. Available from: https://ujms.net/index.php/ujms/article/view/5883

16. Muwanguzi TE, Yadesa TM, Agaba AG. Antibacterial prescription and the associated factors among outpatients diagnosed with respiratory tract infections in Mbarara Municipality, Uganda. BMC Pulm Med. 2021;21(1):1–11.

17. Amuzie CI, Kalu KU, Izuka M, Nwamoh UN, Emma-Ukaegbu U, Odini F, et al. Prevalence, pattern and predictors of self-medication for COVID-19 among residents in Umuahia, Abia State, Southeast Nigeria: policy and public health implications. J Pharm Policy Pract [Internet]. 2022 Dec 2;15(1):34. Available from: https://joppp.biomedcentral.com/articles/10.1186/s40545-022-00429-9

18. Dare SS, Eze ED, Echoru I, Usman IM, Ssempijja F, Bukenya EE, et al. Behavioural Response To Self-Medication Practice Before and During Covid-19 Pandemic in Western Uganda. Patient Prefer Adherence. 2022;16(August):2247–57.

19. John Mathibe L, Perseverance Zwane N. Unnecessary antimicrobial prescribing for upper respiratory tract infections in children in Pietermaritzburg, South Africa. Afr Health Sci [Internet]. 2020 Oct 7;20(3):1133–42. Available from: https://www.ajol.info/index.php/ahs/article/view/200304

20. Odoki M, Aliero AA, Tibyangye J, Nyabayo Maniga J, Wampande E, Kato CD, et al. Prevalence of Bacterial Urinary Tract Infections and Associated Factors among Patients Attending Hospitals in Bushenyi District, Uganda. Int J Microbiol. 2019;

21. Opollo MS, Otim TC, Kizito W, Thekkur P, Kumar AM V., Kitutu FE, et al. Infection Prevention and Control at Lira University Hospital, Uganda: More Needs to Be Done. Trop Med Infect Dis [Internet]. 2021 May 1;6(2):69. Available from: https://www.mdpi.com/2414-6366/6/2/69

22. Mączyńska B, Frej-Mądrzak M, Sarowska J, Woronowicz K, Choroszy-Król I, Jama-Kmiecik A. Evolution of Antibiotic Resistance in Escherichia coli and Klebsiella pneumoniae Clinical Isolates in a Multi-Profile Hospital over 5 Years (2017–2021). J Clin Med [Internet]. 2023 Mar 21;12(6):2414. Available from: https://www.mdpi.com/2077-0383/12/6/2414

23. Shaikh S, Fatima J, Shakil S, Rizvi SMD, Kamal MA. Antibiotic resistance and extended spectrum beta-lactamases: Types, epidemiology and treatment. Saudi J Biol Sci [Internet]. 2015;22(1):90–101. Available from: 10.1016/j.sjbs.2014.08.002

24. Mboowa G, Aruhomukama D, Sserwadda I, Kitutu FE, Davtyan H, Owiti P, et al. Increasing antimicrobial resistance in surgical wards at mulago national referral hospital, uganda, from 2014 to 2018—cause for concern? Trop Med Infect Dis. 2021;

25. Moore CC, Jacob ST, Banura P, Zhang J, Stroup S, Boulware DR, et al. Etiology of Sepsis in Uganda Using a Quantitative Polymerase Chain Reaction-based TaqMan Array Card. Clin Infect Dis [Internet]. 2019 Jan 7;68(2):266–72. Available from: https://academic.oup.com/cid/article/68/2/266/5032704

26. Suvada J, Tumbu P, Iriso R, Russel HN, Namagala E, Kawanguzi P, et al. Bloodstream infections among children living in rural settings in Uganda and predictors of mortality outcome. Int J Infect Dis [Internet]. 2014 Apr;21:104. Available from: https://linkinghub.elsevier.com/retrieve/pii/S1201971214007024

27. Mugerwa I, Nabadda SN, Midega J, Guma C, Kalyesubula S, Muwonge A. Antimicrobial Resistance Situational Analysis 2019–2020: Design and Performance for Human Health Surveillance in Uganda. Trop Med Infect Dis [Internet]. 2021 Sep 29;6(4):178. Available from: https://www.mdpi.com/2414-6366/6/4/178

28. Uganda Ministry of Health. National microbiology reference laboratory (NMRL) Laboratory Handbook. 2020. p. 86.

29. Tomczyk S, Taylor A, Brown A, De Kraker MEA, El-Saed A, Alshamrani M, et al. Impact of the COVID-19 pandemic on the surveillance, prevention and control of antimicrobial resistance: A global survey. Journal of Antimicrobial Chemotherapy. 2021.

30. Frawley T, van Gelderen F, Somanadhan S, Coveney K, Phelan A, Lynam-Loane P, et al. The impact of COVID-19 on health systems, mental health and the potential for nursing. Ir J Psychol Med [Internet]. 2021 Sep 16;38(3):220–6. Available from: https://www.cambridge.org/core/product/identifier/S0790966720001056/type/journal_arti cle

31. Haileamlak A. The impact of COVID-19 on health and health systems. Ethiop J Health Sci [Internet]. 2021 Nov;31(6):1073–4. Available from: http://www.ncbi.nlm.nih.gov/pubmed/35392335

