## Supplementary material for "The epidemiology of antibiotic-resistant clinical pathogens in Uganda": na

Supplementary Material: **The epidemiology of antibiotic-resistant clinical pathogens in Uganda**

Uganda’s sentinel site AMR surveillance system

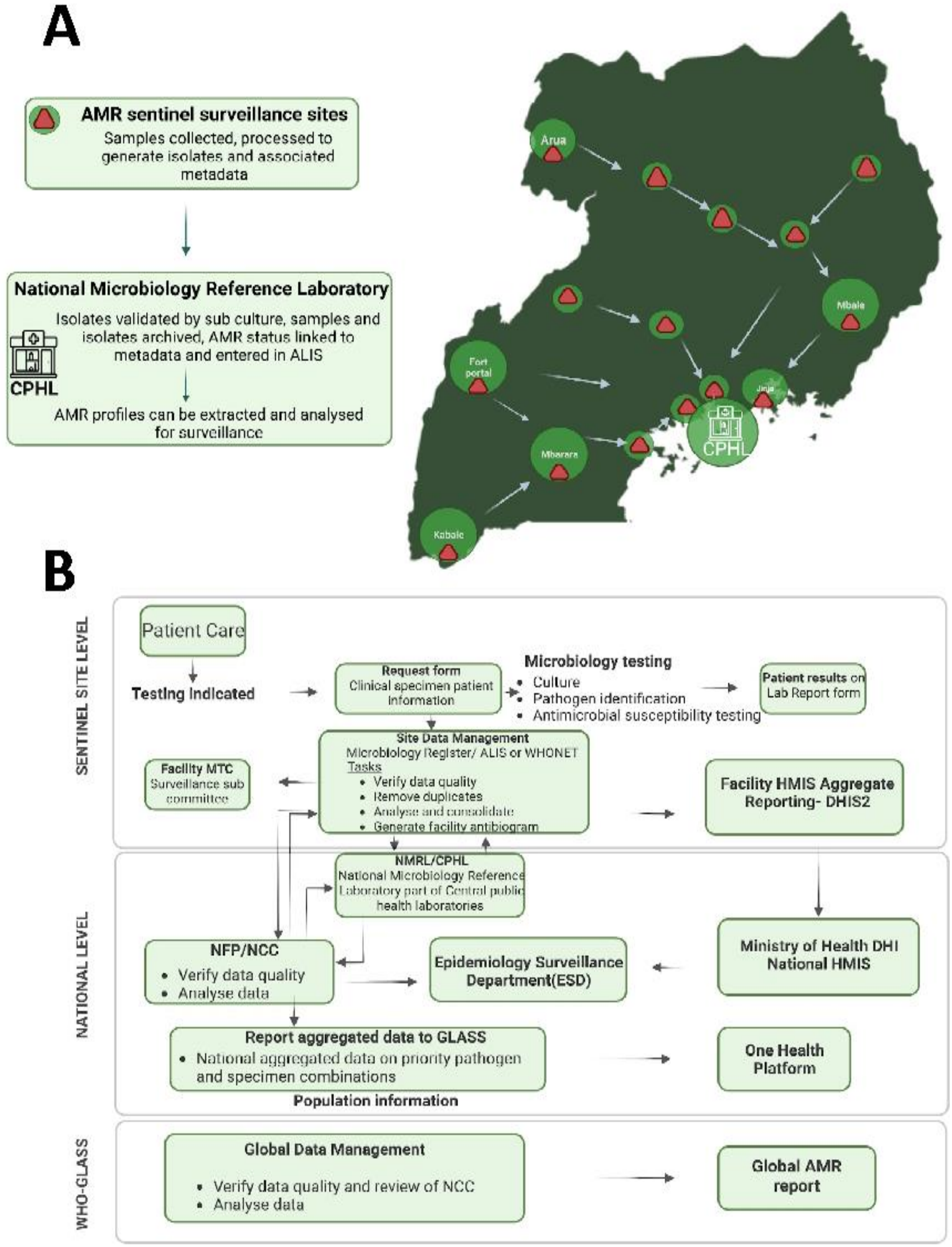

**Fig S1**: **-** The structure of Uganda’s AMR sentinel site surveillance for human health. A- Map of the AMR sentinel surveillance sites and the flow of samples to the National Microbiology Reference Laboratories via the National sample transport network. B- Data flow from AMR sentinel surveillance sites to World health organisation for global reporting. The size of the circle on the map represents the number of isolates collected at a site.

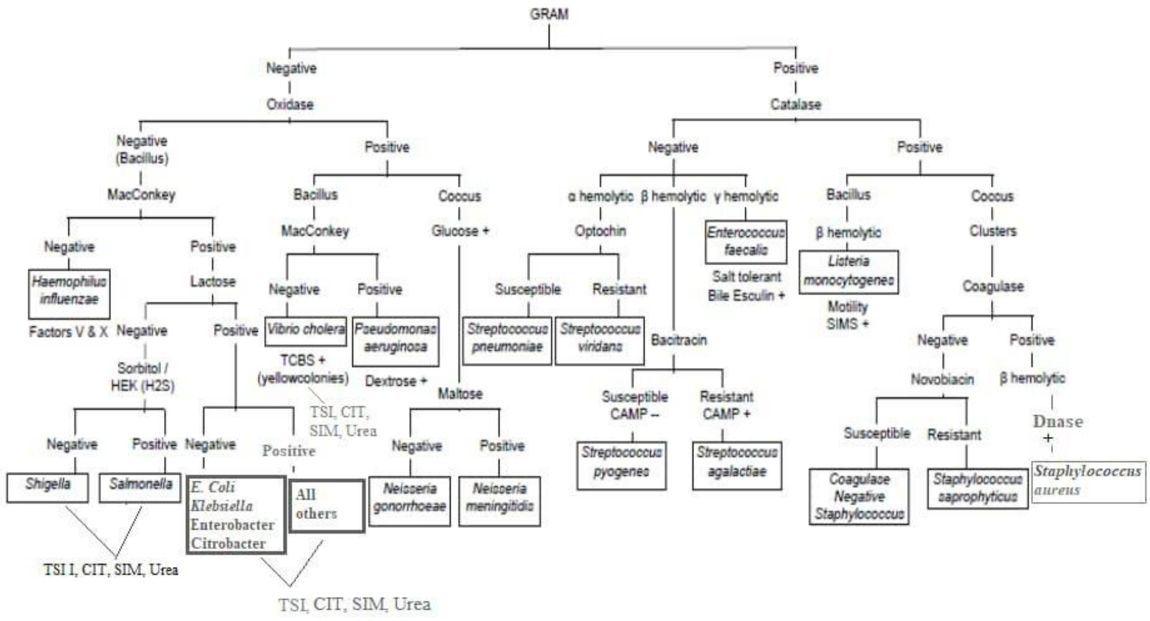

**Fig S2:** Bacteria identification flow chart.

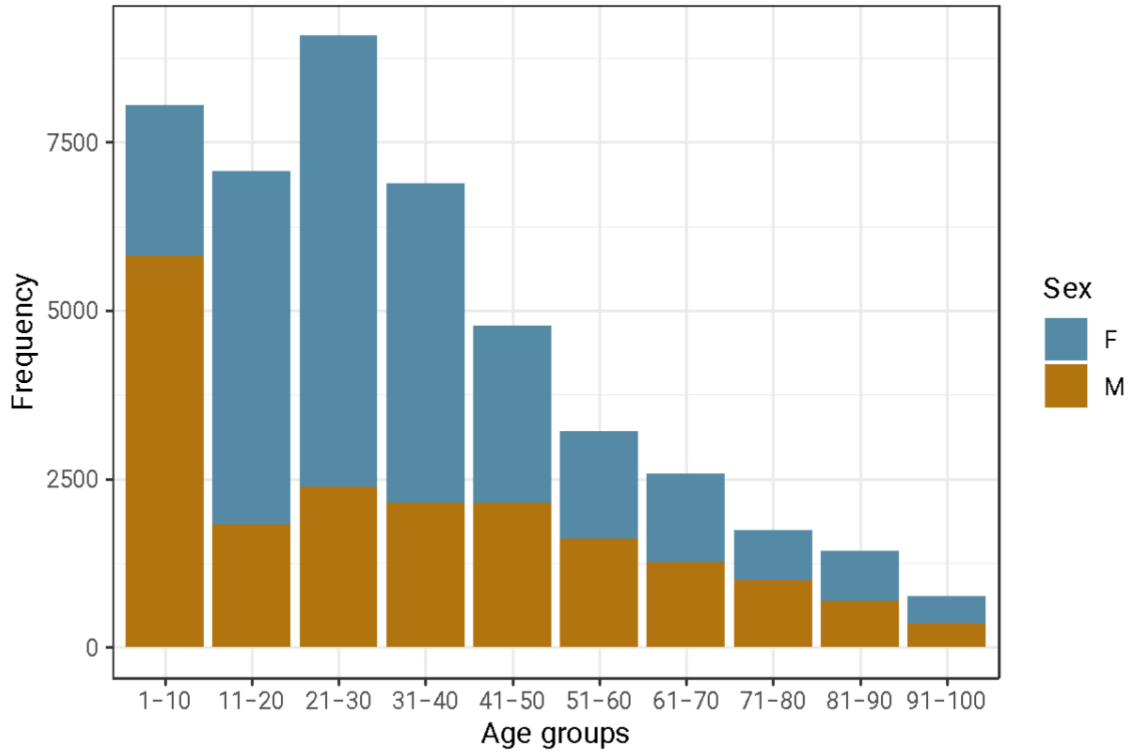

**Figure S3**: The distribution of sentinel surveillance participants by age and gender.

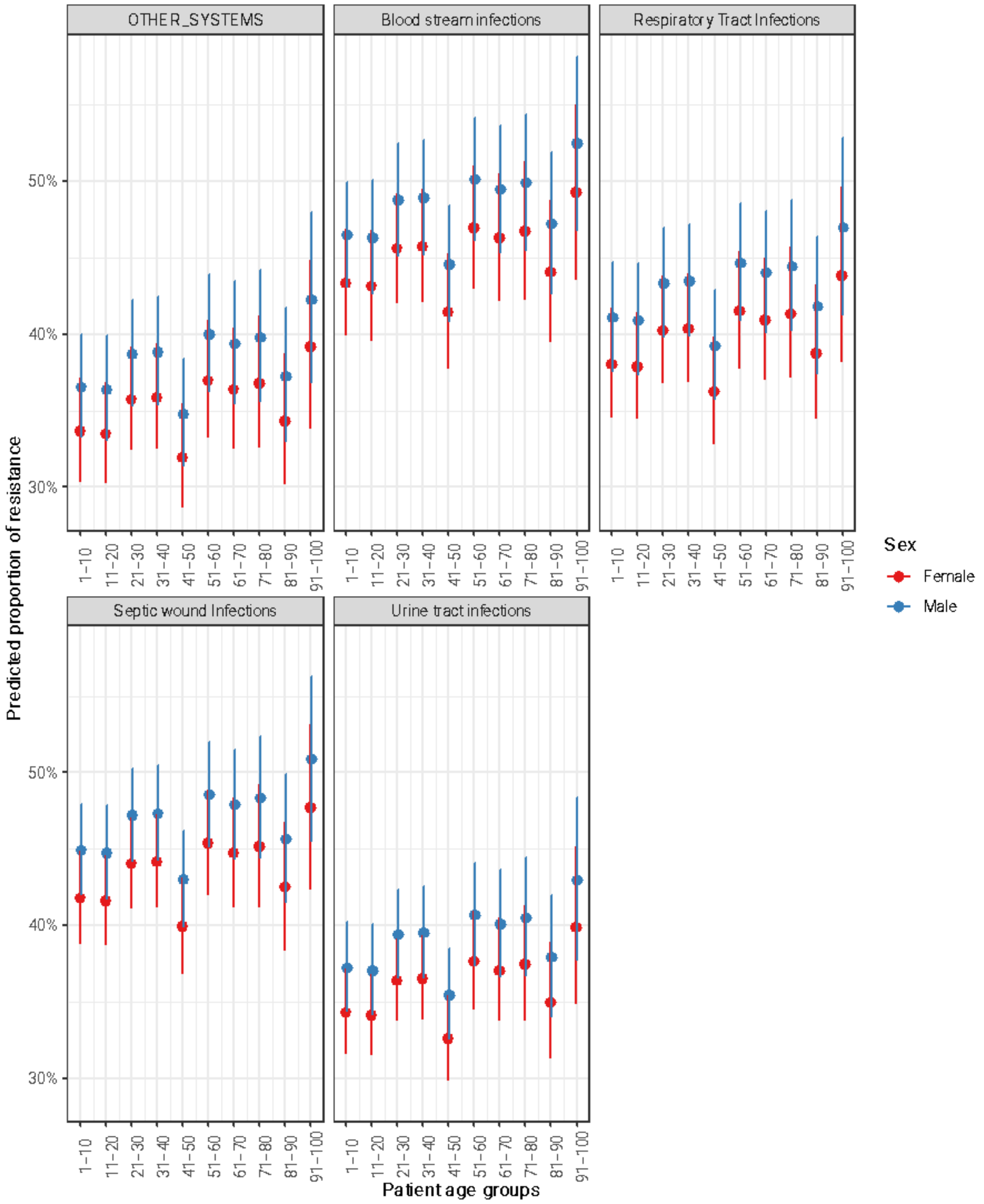

**Figure S4:** Model predicted prevalence of AMR, visualised by age and predicted resistance coloured by gender and faceted by clinical syndromes. The estimated ABR is highest and lowest for BSI and RTIs

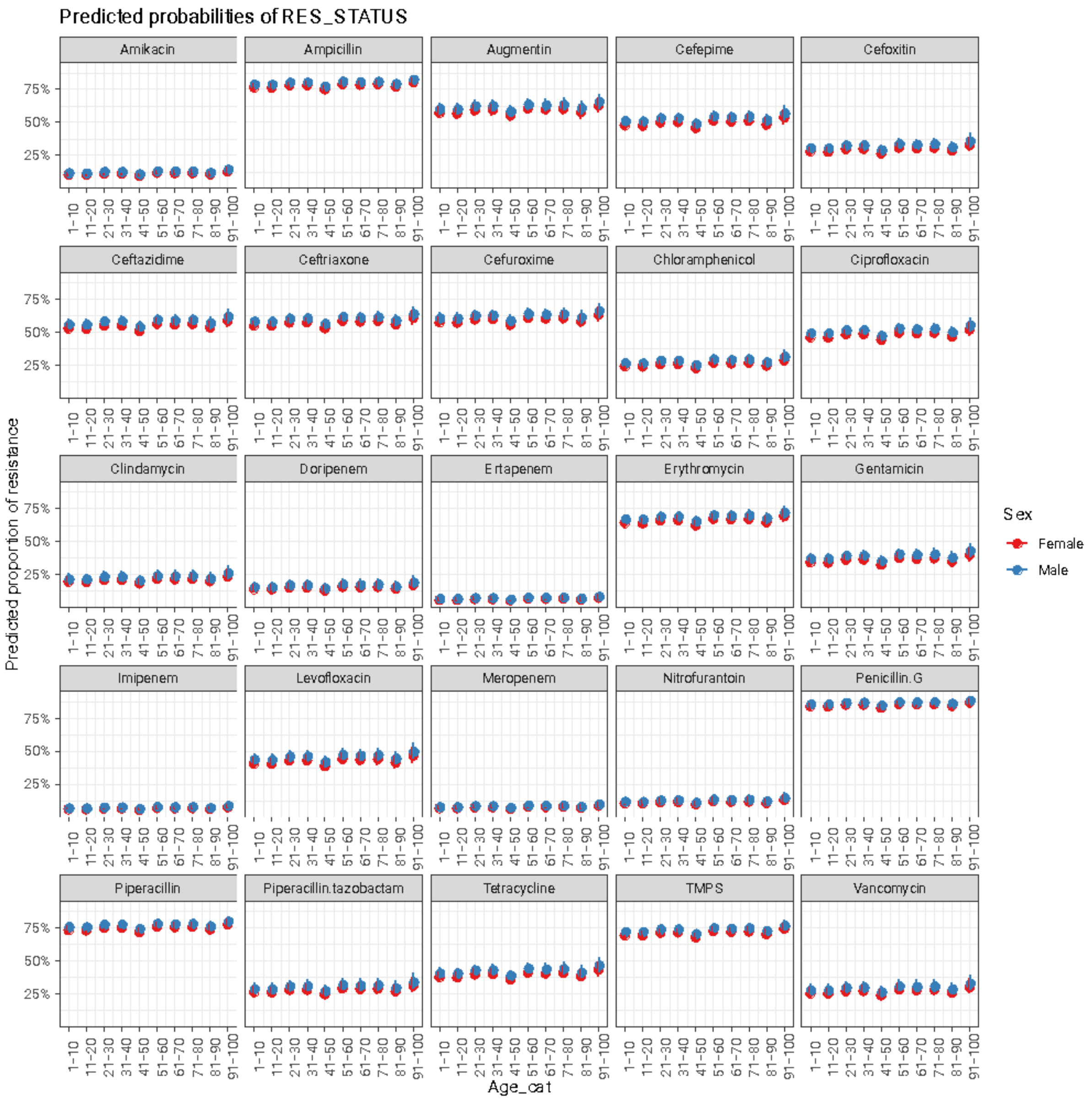

**Figure S5:** Model predicted prevalence of AMR, visualised by age and predicted resistance coloured by gender and faceted by antibiotic. The estimated ABR is highest and lowest for BSI and RTIs

**Table S1**: The descriptive summary of the AMR sentinel surveillance of Uganda characterising the culture recovery rate as a key diagnostics performance indicator. The bold figures represent the highest and lowest CRR measure per category.

| Variable | Levels | Culture recovery | | | Culture Recovery Rate (CRR) (%) |
|---|---|---|---|---|---|
| | | Gram- Negative | Gram- Positive | No Growth | |
| Gender | Male | 1543 | 834 | 1155 | 67.2 |
| | Female | 2015 | 1178 | 2308 | 58.1 |
| Age | 1-10 | 604 | 319 | 269 | 77.5 |
| | 11-20 | 563 | 323 | 349 | 71.7 |
| | 21-30 | 671 | 458 | 985 | 53.4 |
| | 31-40 | 550 | 354 | 987 | **47.8** |
| | 41-50 | 371 | 209 | 415 | 58.2 |
| | 51-60 | 267 | 121 | 199 | 66.0 |
| | 61-70 | 210 | 96 | 109 | 73.7 |
| | 71-80 | 144 | 61 | 76 | 72.9 |
| | 81-90 | 117 | 45 | 51 | 76.0 |
| | 91-100 | 61 | 26 | 23 | **79.0** |
| Region | Eastern | 274 | 196 | 162 | 74.3 |
| | East Central | 165 | 168 | 91 | 78.5 |
| | Central | 508 | 329 | 2095 | **28.5** |
| | West Nile | 177 | 87 | 95 | 73.5 |
| | Western | 1812 | 1097 | 718 | **80.2** |
| | Mid-Western | 591 | 125 | 223 | 76.2 |
| | Northern | 17 | 6 | 51 | 31.0 |
| | North East | 14 | 6 | 28 | 41.6 |
| Year | 2019 | 503 | 220 | 1297 | **35.7** |
| | 2020 | 1833 | 1075 | 1230 | 70.2 |
| | 2021 | 1222 | 717 | 936 | 67.4 |
| Syndrome | RTIs | 305 | 59 | 120 | 75.2 |
| | BSIs | 173 | 278 | 276 | 62.0 |
| | GIT | 21 | 0 | 29 | 42.0 |
| | SWIs | 1279 | 631 | 210 | **90.0** |
| | UTIs | 1543 | 958 | 2279 | 53.2 |
| | Neurological | 14 | 5 | 43 | **30.6** |
| | Others | 223 | 81 | 506 | 37.5 |

**Table S2:** The output from mixed effects logistic regression model, here we compare two models for each of the gram-positive bacteria using the sjplot package in R

| | E. Faecalis | S. aureus |
|---|---|---|

| *Predictors* | *Odds Ratios* | *p* | *Odds Ratios* | *p* |
|---|---|---|---|---|
| (Intercept) | 0.88 (0.66 – 1.16 ) | 0.369 | 0.62 (0.27 – 1.45) | 0.272 |
| Sex [Male] | 0.92 (0.83 – 1.02 ) | 0.126 | 0.92 (0.71 – 1.18) | 0.502 |
| Age_cat11-20 | 1.09 (0.92 – 1.29 ) | 0.330 | 0.63 * (0.41 – 0.96) | **0.033** |
| Age_cat21-30 | 1.26 ** (1.08 – 1.48) | **0.004** | 0.59 * (0.39 – 0.89) | **0.012** |
| Age_cat31-40 | 1.27 ** (1.07 – 1.51) | **0.007** | 0.40 *** (0.26 – 0.60) | **<0.001** |
| Age_cat41-50 | 1.14 (0.93 – 1.39 ) | 0.201 | 0.71 (0.46 – 1.11) | 0.136 |
| Age_cat51-60 | 1.05 (0.84 – 1.30 ) | 0.681 | 0.63 (0.36 – 1.13) | 0.125 |
| Age_cat61-70 | 1.10 (0.87 – 1.41 ) | 0.425 | 1.00 (0.58 – 1.72) | 0.996 |
| Age_cat71-80 | 1.57 ** (1.18 – 2.09) | **0.002** | 0.40 ** (0.20 – 0.80) | **0.010** |
| Age_cat81-90 | 1.25 (0.88 – 1.78 ) | 0.213 | 0.69 (0.33 – 1.45) | 0.328 |
| Age_cat91-100 | 0.52 ** (0.32 – 0.85) | **0.009** | 1.25 (0.62 – 2.54) | 0.539 |
| Syndrome [BSI] | 0.90 (0.74 – 1.10 ) | 0.294 | 1.09 (0.58 – 2.06) | 0.796 |
| Syndrome [RTI] | 0.97 (0.72 – 1.31 ) | 0.852 | 0.44 (0.13 – 1.43) | 0.173 |
| Syndrome [SWI] | 0.58 *** (0.48 – 0.69) | **<0.001** | 0.86 (0.48 – 1.55) | 0.619 |
| Syndrome [UTI] | 0.78 ** (0.65 – 0.94) | **0.007** | 0.65 (0.37 – 1.13) | 0.127 |
| Spectrum [Narrow] | 0.45 *** (0.40 – 0.51) | **<0.001** | 1.83 *** (1.32 – 2.52) | **<0.001** |
| Classification [Watch] | 1.31 *** (1.18 – 1.45) | **<0.001** | | |
| YEAR [2020] | | | 2.24 *** (1.66 – 3.02) | **<0.001** |
| YEAR [2021] | | | 2.53 *** (1.85 – 3.48) | **<0.001** |
| Random Effects | | | | |
| $\sigma^2$ | 3.29 | | 3.29 | |
| $\tau_{00}$ | 0.02 YEAR | | 0.12 Classification | |
| ICC | 0.01 | | 0.04 | |
| N | 3 YEAR | | 2 Classification | |
| Observations | 7279 | | 1982 | |
| Marginal $R^2$ / Conditional $R^2$ | 0.054 / 0.061 | | 0.109 / 0.141 | |
| | | | | * *p<0.05* ** *p<0.01* *** *p<0.001* |

**Table S3**:The output for mixed effects logistic regression model where the outcome was status of phenotypic resistance (resistance or not resistant). Here we compare between four gram negative clinical pathogens using the sjplot package in R.

| | *A.baumanni* | | *C.Fruendii* | | *E.coli* | | *K. Pneumoniae* | |
|---|---|---|---|---|---|---|---|---|
| *Predictors* | *Odds Ratios* | *p* | *Odds Ratios* | *p* | *Odds Ratios* | *p* | *Odds Ratios* | *p* |
| (Intercept) | 1.01 (0.44 – 2.33) | 0.986 | 0.41 ** (0.21 – 0.78) | **0.006** | 0.82 (0.57 – 1.17) | 0.274 | 0.41 (0.16 – 1.07) | 0.069 |
| Sex [Male] | 0.89 (0.67 – 1.18) | 0.409 | 0.72 ** (0.58 – 0.88) | **0.002** | 1.18 *** (1.08 – 1.28) | **<0.001** | 1.25 *** (1.10 – 1.42) | **0.001** |
| Age_cat11-20 | 1.08 (0.75 – 1.57) | 0.674 | 0.99 (0.71 – 1.37) | 0.936 | 0.91 (0.80 – 1.04) | 0.163 | 1.16 (0.94 – 1.43) | 0.175 |
| Age_cat21-30 | 1.16 (0.75 – 1.79) | 0.516 | 1.23 (0.88 – 1.71) | 0.220 | 0.90 (0.79 – 1.02) | 0.097 | 1.29 * (1.06 – 1.59) | **0.013** |
| Age_cat31-40 | 1.07 (0.70 – 1.62) | 0.768 | 0.99 (0.69 – 1.41) | 0.946 | 0.99 (0.87 – 1.13) | 0.861 | 1.63 *** (1.32 – 2.02) | **<0.001** |
| Age_cat41-50 | 0.64 (0.40 – 1.04) | 0.074 | 1.01 (0.70 – 1.47) | 0.948 | 0.78 *** (0.68 – 0.90) | **0.001** | 1.09 (0.86 – 1.39) | 0.483 |
| Age_cat51-60 | 1.58 (0.78 – 3.19) | 0.206 | 1.16 (0.75 – 1.80) | 0.505 | 0.92 (0.79 – 1.08) | 0.323 | 1.53 *** (1.21 – 1.93) | **<0.001** |
| Age_cat61-70 | 0.87 (0.41 – 1.86) | 0.726 | 0.84 (0.56 – 1.25) | 0.389 | 1.08 (0.90 – 1.30) | 0.385 | 1.21 (0.94 – 1.56) | 0.130 |
| Age_cat71-80 | 1.58 (0.53 – 4.69) | 0.413 | 1.07 (0.57 – 1.98) | 0.839 | 0.97 (0.80 – 1.18) | 0.775 | 1.55 ** (1.16 – 2.06) | **0.003** |
| Age_cat81-90 | 1.28 (0.57 – 2.86) | 0.553 | 1.33 (0.80 – 2.21) | 0.268 | 0.92 (0.75 – 1.14) | 0.471 | 1.06 (0.77 – 1.47) | 0.705 |
| Age_cat91-100 | 2.17 (0.65 – 7.22) | 0.205 | 0.66 (0.23 – 1.92) | 0.446 | 1.57 ** (1.18 – 2.11) | **0.002** | 0.84 (0.51 – 1.39) | 0.506 |
| Syndrome [Blood stream infections] | 2.16 (0.98 – 4.77) | 0.056 | 0.79 (0.36 – 1.76) | 0.570 | 1.17 (0.88 – 1.55) | 0.288 | 1.95 *** (1.42 – 2.67) | **<0.001** |
| Syndrome [Respiratory Tract Infections] | 2.38 ** (1.33 – 4.26) | **0.004** | 2.62 *** (1.56 – 4.42) | **<0.001** | 1.29 (0.96 – 1.72) | 0.094 | 0.73 * (0.55 – 0.95) | **0.020** |
| Syndrome [Septic wound Infections] | 3.12 *** (1.94 – 5.02) | **<0.001** | 2.48 *** (1.68 – 3.65) | **<0.001** | 1.64 *** (1.35 – 2.00) | **<0.001** | 1.30 (1.00 – 1.69) | 0.050 |
| Syndrome [Urine tract infections] | 0.52 (0.27 – 1.00) | 0.050 | 1.40 (0.94 – 2.07) | 0.098 | 0.88 (0.72 – 1.07) | 0.195 | 0.57 *** (0.44 – 0.75) | **<0.001** |
| Spectrum [Narrow] | 1.00 (0.70 – 1.43) | 0.999 | 0.77 (0.58 – 1.02) | 0.072 | 0.68 *** (0.61 – 0.76) | **<0.001** | 1.64 *** (1.39 – 1.94) | **<0.001** |
| Bacteria Species [Acinetobacter spp] | 0.52 *** (0.39 – 0.70) | **<0.001** | | | | | | |
| YEAR [2020] | 0.73 (0.53 – 1.01) | 0.059 | 1.46 * (1.05 – 2.04) | **0.025** | 1.23 *** (1.10 – 1.38) | **<0.001** | 1.60 *** (1.31 – 1.94) | **<0.001** |
| YEAR [2021] | 1.04 (0.71 – 1.53) | 0.837 | 1.48 * (1.03 – 2.12) | **0.033** | 1.57 *** (1.40 – 1.77) | **<0.001** | 1.80 *** (1.47 – 2.21) | **<0.001** |
| Random Effects | | | | | | | | |
| $\sigma^2$ | 3.29 | | 3.29 | | 3.29 | | 3.29 | |
| $\tau_{00}$ | 0.15 Classification | | 0.08 Classification | | 0.04 Classification | | 0.41 Classification | |
| ICC | 0.04 | | 0.02 | | 0.01 | | 0.11 | |
| N | 2 Classification | | 2 Classification | | 2 Classification | | 2 Classification | |
| Observations | 1310 | | 1974 | | 13990 | | 5668 | |
| Marginal $R^2$ / Conditional $R^2$ | 0.114 / 0.153 | | 0.043 / 0.065 | | 0.051 / 0.062 | | 0.072 / 0.175 | |
| | | | | | | | | * *p<0.05* ** *p<0.01* *** *p<0.001* |

**Table S4. Summary of culture recovery rate across the 3 years**

| *Organism* | *Years* | | | ***Total*** |
|---|---|---|---|---|
| | 2019 | 2020 | 2021 | |
| *Acinetobacter baumannii* | 11 (20 %) | 34 (61.8 %) | 10 (18.2 %) | 55 (100 %) |
| *Acinetobacter* spp | 36 (19.9 %) | 92 (50.8 %) | 53 (29.3 %) | 181 (100 %) |
| *Beta- Hemolytic streptococcus* | 12 (10.6 %) | 67 (59.3 %) | 34 (30.1 %) | 113 (100 %) |
| *Candida albicans* | 159 (31.1 %) | 210 (41.1 %) | 142 (27.8 %) | 511 (100 %) |
| *Candida* spp | 46 (22.2 %) | 92 (44.4 %) | 69 (33.3 %) | 207 (100 %) |
| *Citrobacter freundii* | 24 (10.5 %) | 139 (61 %) | 65 (28.5 %) | 228 (100 %) |
| *Citrobacter* spp | 3 (5.6 %) | 29 (53.7 %) | 22 (40.7 %) | 54 (100 %) |
| Coagulase-negative *Staphylococcus* | 82 (9.7 %) | 417 (49.5 %) | 344 (40.8 %) | 843 (100 %) |
| *Cryptococcus* spp | 2 (11.8 %) | 8 (47.1 %) | 7 (41.2 %) | 17 (100 %) |
| *Enterobacter* spp | 14 (9.9 %) | 63 (44.7 %) | 64 (45.4 %) | 141 (100 %) |
| *Enterococcus faecalis* | 64 (14.3 %) | 244 (54.5 %) | 140 (31.2 %) | 448 (100 %) |
| Enterococcus spp | 7 (18.4 %) | 14 (36.8 %) | 17 (44.7 %) | 38 100 %) |
| *Escherichia coli* | 205 (12.5 %) | 849 (51.8 %) | 584 (35.7 %) | 1638 (100 %) |
| *Klebsiella pneumoniae* | 62 (10.1 %) | 336 (54.9 %) | 214 (35 %) | 612 (100 %) |
| *Klebsiella* spp | 15 (18.3 %) | 39 (47.6 %) | 28 (34.1 %) | 82 (100 %) |

| | | | | |
|---|---|---|---|---|
| Mixed Bacterial Growth | 8<br>(44.4 %) | 4<br>(22.2 %) | 6<br>(33.3 %) | 18<br>(100 %) |
| *Morganella morganii* | 3<br>(12.5 %) | 13<br>(54.2 %) | 8<br>(33.3 %) | 24<br>(100 %) |
| *Neisseria gonorrhoeae* | 0<br>(0 %) | 0<br>(0 %) | 34<br>(100 %) | 34<br>(100 %) |
| No growth | 1065<br>(41.1 %) | 849<br>(32.8 %) | 677<br>(26.1 %) | 2591<br>(100 %) |
| *Proteus* spp | 43<br>(26.1 %) | 73<br>(44.2 %) | 49<br>(29.7 %) | 165<br>(100 %) |
| *Providencia* spp | 0<br>(0 %) | 13<br>(59.1 %) | 9<br>(40.9 %) | 22<br>(100 %) |
| *Pseudomonas aeruginosa* | 31<br>(20.7 %) | 74<br>(49.3 %) | 45<br>(30 %) | 150<br>(100 %) |
| *Pseudomonas* spp | 4<br>(14.3 %) | 11<br>(39.3 %) | 13<br>(46.4 %) | 28<br>(100 %) |
| *Salmonella* spp | 19<br>(30.6 %) | 36<br>(58.1 %) | 7<br>(11.3 %) | 62<br>(100 %) |
| *Serratia* spp | 3<br>(21.4 %) | 5<br>(35.7 %) | 6<br>(42.9 %) | 14<br>(100 %) |
| *Shigella* spp | 6<br>(35.3 %) | 11<br>(64.7 %) | 0<br>(0 %) | 17<br>(100 %) |
| *Staphylococcus aureus* | 57<br>(8.5 %) | 396<br>(59.4 %) | 214<br>(32.1 %) | 667<br>(100 %) |
| *Staphylococcus saprophyticus* | 6<br>(75 %) | 2<br>(25 %) | 0<br>(0 %) | 8<br>(100 %) |
| *Streptococcus pneumoniae* | 3<br>(42.9 %) | 2<br>(28.6 %) | 2<br>(28.6 %) | 7<br>(100 %) |
| *Vibrio cholerae* | 22<br>(44.9 %) | 16<br>(32.7 %) | 11<br>(22.4 %) | 49<br>(100 %) |
| **Total** | **2012<br>(22.3 %)** | **4138<br>(45.9 %)** | **2874<br>(31.8 %)** | **9024<br>(100 %)** |

*$\chi^2$=1108.281 · df=58 · Cramer's V=0.248 · Fisher's p=0.000*

**Table S5**:The output for mixed effects logistic regression model where the outcome was status of phenotypic resistance (resistance or not resistant). Here we compare between four major antibiotic classes using the sjplot package in R

| | Beta lactams | | Penicillin | | Cephalosporins | | Carbapenems | |
|---|---|---|---|---|---|---|---|---|
| *Predictors* | *Odds Ratios* | *p* | *Odds Ratios* | *p* | *Odds Ratios* | *p* | *Odds Ratios* | *p* |
| (Intercept) | 0.48 (0.18 – 1.28) | 0.144 | 5.17 * (1.09 – 24.59) | **0.039** | 0.38 * (0.16 – 0.87) | **0.023** | 0.05 *** (0.02 – 0.10) | **<0.001** |
| Sex [Male] | 0.74 * (0.56 – 0.97) | **0.031** | 0.93 (0.77 – 1.12) | 0.435 | 1.19 ** (1.05 – 1.34) | **0.006** | 1.20 (0.96 – 1.51) | 0.117 |
| YEAR [2020] | 5.68 *** (3.95 – 8.17) | **<0.001** | 1.80 *** (1.40 – 2.31) | **<0.001** | 1.28 ** (1.07 – 1.53) | **0.006** | 1.19 (0.86 – 1.65) | 0.301 |
| YEAR [2021] | 1.79 ** (1.25 – 2.55) | **0.001** | 2.14 *** (1.63 – 2.81) | **<0.001** | 2.27 *** (1.88 – 2.75) | **<0.001** | 1.31 (0.92 – 1.85) | 0.132 |
| Age_cat11-20 | 0.86 (0.55 – 1.35) | 0.513 | 1.05 (0.77 – 1.43) | 0.778 | 0.80 * (0.66 – 0.97) | **0.024** | 1.37 (0.96 – 1.94) | 0.079 |
| Age_cat21-30 | 0.93 (0.62 – 1.39) | 0.713 | 1.14 (0.85 – 1.53) | 0.384 | 0.80 * (0.67 – 0.97) | **0.024** | 1.00 (0.70 – 1.44) | 0.985 |
| Age_cat31-40 | 1.47 (0.95 – 2.27) | 0.087 | 0.95 (0.70 – 1.28) | 0.724 | 0.84 (0.69 – 1.03) | 0.087 | 1.07 (0.74 – 1.56) | 0.721 |
| Age_cat41-50 | 1.09 (0.68 – 1.73) | 0.723 | 1.12 (0.80 – 1.57) | 0.509 | 0.61 *** (0.49 – 0.76) | **<0.001** | 0.90 (0.60 – 1.35) | 0.625 |
| Age_cat51-60 | 1.65 * (1.00 – 2.71) | **0.049** | 1.13 (0.77 – 1.64) | 0.541 | 0.90 (0.70 – 1.15) | 0.390 | 1.09 (0.71 – 1.66) | 0.705 |
| Age_cat61-70 | 1.24 (0.69 – 2.23) | 0.472 | 1.70 * (1.08 – 2.67) | **0.023** | 0.84 (0.65 – 1.08) | 0.173 | 1.06 (0.67 – 1.69) | 0.807 |
| Age_cat71-80 | 1.63 (0.85 – 3.13) | 0.141 | 2.02 ** (1.22 – 3.32) | **0.006** | 0.86 (0.63 – 1.17) | 0.341 | 0.73 (0.40 – 1.33) | 0.303 |
| Age_cat81-90 | 1.62 (0.82 – 3.22) | 0.167 | 1.13 (0.69 – 1.86) | 0.625 | 0.69 * (0.50 – 0.95) | **0.022** | 1.34 (0.77 – 2.35) | 0.301 |
| Age_cat91-100 | 2.21 (0.75 – 6.47) | 0.149 | 1.92 (0.88 – 4.21) | 0.103 | 1.23 (0.78 – 1.96) | 0.371 | 1.81 (0.87 – 3.77) | 0.110 |
| Syndrome [Blood stream infections] | 1.42 (0.65 – 3.09) | 0.380 | 2.96 *** (1.85 – 4.72) | **<0.001** | 2.15 *** (1.55 – 2.96) | **<0.001** | 1.62 (0.82 – 3.18) | 0.165 |
| Syndrome [Respiratory Tract Infections] | 1.14 (0.59 – 2.21) | 0.700 | 0.93 (0.61 – 1.41) | 0.738 | 1.65 *** (1.23 – 2.22) | **0.001** | 1.86 * (1.05 – 3.30) | **0.034** |
| Syndrome [Septic wound Infections] | 0.85 (0.47 – 1.53) | 0.592 | 1.84 *** (1.31 – 2.58) | **<0.001** | 2.52 *** (1.98 – 3.20) | **<0.001** | 2.40 *** (1.48 – 3.91) | **<0.001** |
| Syndrome [Urine tract infections] | 0.66 (0.37 – 1.20) | 0.173 | 0.91 (0.65 – 1.26) | 0.555 | 1.37 * (1.07 – 1.75) | **0.012** | 1.07 (0.65 – 1.78) | 0.787 |

| | | | | | | | |
|---|---|---|---|---|---|---|---|
| Gram status [Gram Positive] | | | 0.05 *** (0.04 – 0.07) | **<0.001** | 4.14 *** (3.29 – 5.21) | **<0.001** | |
| Random Effects | | | | | | | |
| $\sigma^2$ | 3.29 | | 3.29 | | 3.29 | | 3.29 |
| $\tau_{00}$ | 0.25 $_{\text{Antibiotics}}$ | | 1.74 $_{\text{Antibiotics}}$ | | 0.63 $_{\text{Antibiotics}}$ | | 0.16 $_{\text{Antibiotics}}$ |
| ICC | 0.07 | | 0.35 | | 0.16 | | 0.05 |
| N | 2 $_{\text{Antibiotics}}$ | | 3 $_{\text{Antibiotics}}$ | | 4 $_{\text{Antibiotics}}$ | | 4 $_{\text{Antibiotics}}$ |
| Observations | 1393 | | 4134 | | 6129 | | 4601 |
| Marginal $R^2$ / Conditional $R^2$ | 0.140 / 0.200 | | 0.298 / 0.541 | | 0.103 / 0.246 | | 0.054 / 0.100 |
| * *p<0.05* ** *p<0.01* *** *p<0.001* | | | | | | | |

**Table S6**: The output for mixed effects logistic regression model where the outcome was status of phenotypic resistance (resistance or not resistant). Here we compare between other antibiotic classes using the sjplot package in R. Here in some of the classes we had one antibiotic represented.

| | Gentamycin | | Erythromycin | | TMPS | | Fluoroquinolones | | Tetracyclines | |
|---|---|---|---|---|---|---|---|---|---|---|
| *Predictors* | *Odds Ratios* | *p* | *Odds Ratios* | *p* | *Odds Ratios* | *p* | *Odds Ratios* | *p* | *Odds Ratios* | *p* |
| (Intercept) | 0.33 ** (0.16 – 0.67) | **0.002** | 0.94 (0.54 – 1.63) | 0.833 | 2.20 ** (1.35 – 3.58) | **0.001** | 1.05 (0.71 – 1.56) | 0.797 | 0.04 * (0.00 – 0.98) | **0.049** |
| Sex [Male] | 1.00 (0.80 – 1.23) | 0.969 | 0.88 (0.70 – 1.12) | 0.293 | 1.07 (0.89 – 1.29) | 0.485 | 1.28 *** (1.12 – 1.48) | **<0.001** | 1.29 (0.91 – 1.82) | 0.157 |
| YEAR [2020] | 1.46 ** (1.10 – 1.94) | **0.009** | | | 0.80 (0.60 – 1.06) | 0.120 | | | 1.16 (0.70 – 1.90) | 0.565 |
| YEAR [2021] | 1.44 * (1.07 – 1.95) | **0.017** | | | 1.31 (0.96 – 1.79) | 0.084 | | | 1.73 * (1.07 – 2.79) | **0.024** |
| Age_cat11-20 | 0.71 * (0.51 – 0.98) | **0.040** | 1.06 (0.72 – 1.58) | 0.757 | 1.16 (0.87 – 1.55) | 0.304 | | | 0.82 (0.48 – 1.39) | 0.455 |
| Age_cat21-30 | 0.69 * (0.50 – 0.94) | **0.020** | 1.81 ** (1.24 – 2.63) | **0.002** | 1.13 (0.85 – 1.50) | 0.385 | | | 1.15 (0.68 – 1.96) | 0.593 |
| Age_cat31-40 | 0.84 (0.61 – 1.17) | 0.298 | 1.46 (0.98 – 2.18) | 0.065 | 1.20 (0.88 – 1.62) | 0.245 | | | 1.29 (0.72 – 2.30) | 0.394 |
| Age_cat41-50 | 0.59 ** (0.40 – 0.87) | **0.008** | 1.41 (0.90 – 2.21) | 0.136 | 0.93 (0.67 – 1.27) | 0.637 | | | 0.83 (0.48 – 1.44) | 0.515 |
| Age_cat51-60 | 0.75 (0.49 – 1.17) | 0.205 | 1.36 (0.82 – 2.26) | 0.234 | 1.40 (0.95 – 2.04) | 0.087 | | | 1.63 (0.80 – 3.31) | 0.177 |
| Age_cat61-70 | 0.73 (0.47 – 1.15) | 0.179 | 1.13 (0.66 – 1.93) | 0.659 | 1.14 (0.76 – 1.71) | 0.518 | | | 1.45 (0.63 – 3.34) | 0.381 |
| Age_cat71-80 | 0.52 * (0.30 – 0.90) | **0.019** | 1.83 (0.93 – 3.59) | 0.079 | 0.99 (0.62 – 1.58) | 0.961 | | | 1.51 (0.63 – 3.60) | 0.358 |
| Age_cat81-90 | 0.44 * (0.23 – 0.84) | **0.013** | 1.84 (0.81 – 4.19) | 0.146 | 1.40 (0.81 – 2.42) | 0.230 | | | 1.16 (0.53 – 2.55) | 0.715 |
| Age_cat91-100 | 0.64 (0.29 – 1.41) | 0.270 | 0.59 (0.24 – 1.47) | 0.256 | 0.58 (0.29 – 1.16) | 0.124 | | | 1.12 (0.37 – 3.38) | 0.836 |
| Syndrome [BSI] | 2.79 *** (1.67 – 4.67) | **<0.001** | 1.24 (0.79 – 1.94) | 0.344 | 1.06 (0.70 – 1.60) | 0.785 | 1.23 (0.88 – 1.74) | 0.228 | 3.12 * (1.17 – 8.31) | **0.023** |
| Syndrome [RTI | 1.64 (0.98 – 2.75) | 0.059 | 0.96 (0.50 – 1.83) | 0.904 | 0.60 * (0.38 – 0.92) | **0.021** | 1.18 (0.82 – 1.69) | 0.364 | 1.35 (0.61 – 3.01) | 0.463 |
| Syndrome SWI] | 2.00 ** (1.31 – 3.04) | **0.001** | 0.55 ** (0.37 – 0.81) | **0.003** | 0.91 (0.65 – 1.27) | 0.581 | 1.56 ** (1.19 – 2.05) | **0.001** | 3.68 *** (1.87 – 7.25) | **<0.001** |
| Syndrome [UTI] | 0.98 (0.64 – 1.51) | 0.945 | 1.43 (0.97 – 2.12) | 0.071 | 1.19 (0.85 – 1.67) | 0.318 | 1.23 (0.92 – 1.62) | 0.157 | 4.11 *** (2.06 – 8.20) | **<0.001** |
| Genus [Citrobacter] | | | | | | | 0.52 ** (0.35 – 0.78) | **0.002** | | |

| | | | | | | | | | |
|---|---|---|---|---|---|---|---|---|---|
| Genus [Enterobacter] | | | | | | | 0.44 ** (0.26 – 0.73) | **0.002** | |
| Genus [Enterococcus] | | | | | | | 0.52 ** (0.35 – 0.78) | **0.001** | |
| Genus [Escherichia] | | | | | | | 1.11 (0.81 – 1.53) | 0.513 | |
| Genus [Klebsiella] | | | | | | | 0.34 *** (0.24 – 0.48) | **<0.001** | |
| Genus [Other_genera] | | | | | | | 0.17 *** (0.10 – 0.30) | **<0.001** | |
| Genus [Proteus] | | | | | | | 0.37 *** (0.24 – 0.59) | **<0.001** | |
| Genus [Pseudomonas] | | | | | | | 0.18 *** (0.11 – 0.28) | **<0.001** | |
| Genus [Staphylococcus] | | | | | | | 0.38 *** (0.28 – 0.53) | **<0.001** | |
| Genus [Streptococcus] | | | | | | | 0.07 * (0.01 – 0.56) | **0.012** | |
| Random Effects | | | | | | | | | |
| $\sigma^2$ | 3.29 | | 3.29 | | 3.29 | | 3.29 | | 3.29 |
| $\tau_{00}$ | 0.11 Gram_status | | 0.06 YEAR | | 0.02 Gram_status | | 0.00 Antibiotics | | 5.29 Gram_status |
| ICC | 0.03 | | 0.02 | | 0.01 | | | | 0.62 |
| N | 2 Gram_status | | 3 YEAR | | 2 Gram_status | | 2 Antibiotics | | 2 Gram_status |
| Observations | 2031 | | 1423 | | 2848 | | 4212 | | 1270 |
| Marginal $R^2$ / Conditional $R^2$ | 0.064 / 0.093 | | 0.078 / 0.094 | | 0.029 / 0.034 | | 0.107 / NA | | 0.026 / 0.627 |
| * *p<0.05* ** *p<0.01* *** *p<0.001* | | | | | | | | | |

**Table S7**:The output for mixed effects logistic regression model where the outcome was status of phenotypic resistance (resistance or not resistant). Here we compare between clinical syndromes using the sjplot package in R

| | RTIs | | SWIs | | UTIs | | BSI | | Others | |
|---|---|---|---|---|---|---|---|---|---|---|
| *Predictors* | *Odds Ratios* | *p* | *Odds Ratios* | *p* | *Odds Ratios* | *p* | *Odds Ratios* | *p* | *Odds Ratios* | *p* |
| (Intercept) | 0.10 *** (0.05 – 0.21) | **<0.001** | 0.19 *** (0.13 – 0.28) | **<0.001** | 0.04 *** (0.02 – 0.08) | **<0.001** | 1.06 (0.51 – 2.19) | 0.881 | 0.57 (0.30 – 1.11) | 0.098 |
| Male | 1.53 *** (1.27 – 1.84) | **<0.001** | 0.97 (0.89 – 1.07) | 0.549 | 1.36 *** (1.24 – 1.49) | **<0.001** | 1.10 (0.93 – 1.29) | 0.265 | 0.86 (0.72 – 1.04) | 0.127 |
| Citrobacter | 0.85 (0.51 – 1.41) | 0.527 | 0.48 *** (0.38 – 0.59) | **<0.001** | 2.79 *** (1.74 – 4.46) | **<0.001** | 0.31 * (0.12 – 0.80) | **0.015** | 0.89 (0.51 – 1.55) | 0.669 |
| Enterobacter | 0.35 *** (0.21 – 0.58) | **<0.001** | 1.12 (0.83 – 1.51) | 0.469 | 2.28 ** (1.38 – 3.77) | **0.001** | 0.71 (0.33 – 1.51) | 0.367 | 1.13 (0.62 – 2.06) | 0.689 |
| Enterococcus | 0.28 * (0.08 – 0.98) | **0.046** | 0.36 *** (0.26 – 0.49) | **<0.001** | 1.37 (0.86 – 2.18) | 0.190 | 1.19 (0.60 – 2.39) | 0.620 | 1.98 * (1.02 – 3.83) | **0.043** |
| Escherichia] | 0.92 (0.61 – 1.39) | 0.692 | 0.81 * (0.68 – 0.96) | **0.017** | 3.09 *** (1.98 – 4.81) | **<0.001** | 1.04 (0.53 – 2.02) | 0.915 | 1.92 ** (1.20 – 3.07) | **0.006** |
| Klebsiella] | 0.61 ** (0.43 – 0.87) | **0.006** | 0.67 *** (0.55 – 0.81) | **<0.001** | 2.16 *** (1.37 – 3.39) | **0.001** | 1.41 (0.74 – 2.70) | 0.296 | 2.05 ** (1.26 – 3.33) | **0.004** |
| Other_genera | 0.87 (0.36 – 2.10) | 0.754 | 0.71 * (0.51 – 0.97) | **0.033** | 3.69 *** (1.96 – 6.96) | **<0.001** | 0.43 * (0.21 – 0.88) | **0.021** | 0.29 *** (0.16 – 0.53) | **<0.001** |
| Proteus | 0.57 (0.27 – 1.18) | 0.129 | 0.33 *** (0.26 – 0.40) | **<0.001** | 2.50 ** (1.38 – 4.53) | **0.003** | 0.19 * (0.04 – 0.99) | **0.048** | 0.36 ** (0.19 – 0.69) | **0.002** |
| Pseudomonas | 0.16 *** (0.09 – 0.28) | **<0.001** | 0.13 *** (0.09 – 0.18) | **<0.001** | 1.44 (0.79 – 2.63) | 0.234 | 0.20 ** (0.06 – 0.64) | **0.007** | 0.42 ** (0.23 – 0.78) | **0.005** |
| Staphylococcus | 0.74 (0.46 – 1.21) | 0.236 | 0.21 *** (0.17 – 0.26) | **<0.001** | 2.15 *** (1.36 – 3.40) | **0.001** | 0.59 (0.31 – 1.11) | 0.103 | 1.71 * (1.08 – 2.71) | **0.023** |
| Streptococcus | 0.31 *** (0.15 – 0.62) | **0.001** | 0.11 *** (0.05 – 0.23) | **<0.001** | 0.64 (0.35 – 1.15) | 0.137 | 0.60 (0.25 – 1.44) | 0.254 | 0.70 (0.26 – 1.89) | 0.479 |
| Age 11-20 | 0.89 (0.54 – 1.47) | 0.651 | 1.21 ** (1.05 – 1.39) | **0.007** | 0.78 ** (0.66 – 0.92) | **0.003** | 0.81 (0.62 – 1.07) | 0.143 | 0.58 *** (0.43 – 0.78) | **<0.001** |
| Age 21-30 | 1.01 (0.65 – 1.56) | 0.973 | 1.20 ** (1.05 – 1.38) | **0.009** | 0.86 (0.74 – 1.00) | 0.055 | 0.97 (0.75 – 1.25) | 0.823 | 0.71 * (0.53 – 0.96) | **0.027** |
| Age 31-40 | 1.93 ** (1.24 – 3.00) | **0.003** | 1.17 * (1.01 – 1.36) | **0.034** | 0.85 * (0.72 – 1.00) | **0.047** | 0.76 (0.55 – 1.05) | 0.099 | 0.73 * (0.55 – 0.99) | **0.042** |
| Age 41-50 | 0.92 (0.61 – 1.39) | 0.693 | 0.96 (0.82 – 1.13) | 0.647 | 0.74 *** (0.63 – 0.88) | **0.001** | 0.44 *** (0.30 – 0.65) | **<0.001** | 0.75 (0.52 – 1.07) | 0.114 |
| Age 51-60 | 1.13 (0.74 – 1.73) | 0.571 | 1.08 (0.91 – 1.28) | 0.380 | 1.11 (0.91 – 1.35) | 0.313 | 1.01 (0.71 – 1.44) | 0.964 | 0.53 *** (0.36 – 0.77) | **0.001** |
| Age 61-70 | 0.67 (0.43 – 1.05) | 0.078 | 1.05 (0.88 – 1.27) | 0.580 | 1.12 (0.91 – 1.38) | 0.293 | 1.88 ** (1.21 – 2.92) | **0.005** | 0.90 (0.57 – 1.43) | 0.654 |
| Age 71-80 | 1.21 (0.77 – 1.90) | 0.407 | 0.80 (0.63 – 1.02) | 0.074 | 1.19 (0.94 – 1.50) | 0.153 | 0.74 (0.45 – 1.22) | 0.241 | 0.93 (0.55 – 1.56) | 0.777 |
| Age 81-90 | 1.36 (0.84 – 2.23) | 0.215 | 0.63 ** (0.45 – 0.87) | **0.005** | 0.80 (0.63 – 1.00) | 0.054 | 0.79 (0.50 – 1.25) | 0.318 | 1.24 (0.76 – 2.03) | 0.392 |

| | | | | | | | | | | |
|---|---|---|---|---|---|---|---|---|---|---|
| Age 91-100 | 2.90 $^{*}$<br>(1.02 – 8.19) | **0.045** | 0.96<br>(0.66 – 1.37) | 0.808 | 1.72 $^{**}$<br>(1.23 – 2.41) | **0.001** | 0.53 $^{*}$<br>(0.32 – 0.87) | **0.013** | 0.71<br>(0.40 – 1.26) | 0.246 |
| Ampicillin | 53.35 $^{***}$<br>(25.62 – 111.08) | **<0.001** | 35.50 $^{***}$<br>(25.76 – 48.92) | **<0.001** | 22.27 $^{***}$<br>(17.20 – 28.85) | **<0.001** | | | | |
| Augmentin | 18.80 $^{***}$<br>(9.89 – 35.75) | **<0.001** | 8.15 $^{***}$<br>(5.79 – 11.48) | **<0.001** | 10.97 $^{***}$<br>(8.18 – 14.72) | **<0.001** | | | | |
| Cefepime | 6.60 $^{***}$<br>(3.86 – 11.29) | **<0.001** | 11.30 $^{***}$<br>(8.68 – 14.70) | **<0.001** | 6.09 $^{***}$<br>(4.71 – 7.87) | **<0.001** | | | | |
| Cefoxitin | 2.77 $^{**}$<br>(1.49 – 5.15) | **0.001** | 3.94 $^{***}$<br>(3.01 – 5.15) | **<0.001** | 2.65 $^{***}$<br>(2.00 – 3.50) | **<0.001** | | | | |
| Ceftazidime | 9.13 $^{***}$<br>(5.20 – 16.01) | **<0.001** | 15.03 $^{***}$<br>(11.51 – 19.63) | **<0.001** | 7.24 $^{***}$<br>(5.57 – 9.40) | **<0.001** | | | | |
| Ceftriaxone | 10.30 $^{***}$<br>(6.16 – 17.24) | **<0.001** | 18.15 $^{***}$<br>(14.09 – 23.38) | **<0.001** | 7.30 $^{***}$<br>(5.72 – 9.30) | **<0.001** | | | | |
| Cefuroxime | 9.68 $^{***}$<br>(5.16 – 18.15) | **<0.001** | 23.03 $^{***}$<br>(16.66 – 31.86) | **<0.001** | 7.36 $^{***}$<br>(5.54 – 9.78) | **<0.001** | | | | |
| Chloramphenicol | 2.41 $^{**}$<br>(1.40 – 4.16) | **0.001** | 3.06 $^{***}$<br>(2.38 – 3.94) | **<0.001** | 2.08 $^{***}$<br>(1.61 – 2.68) | **<0.001** | | | | |
| Ciprofloxacin | 5.26 $^{***}$<br>(3.10 – 8.95) | **<0.001** | 8.27 $^{***}$<br>(6.50 – 10.51) | **<0.001** | 7.87 $^{***}$<br>(6.19 – 10.00) | **<0.001** | | | | |
| Clindamycin | 1.91<br>(0.77 – 4.73) | 0.162 | 2.63 $^{***}$<br>(1.79 – 3.86) | **<0.001** | 2.62 $^{***}$<br>(1.80 – 3.83) | **<0.001** | | | | |
| Doripenem | 1.92<br>(0.80 – 4.61) | 0.145 | 1.77 $^{**}$<br>(1.21 – 2.60) | **0.003** | 1.20<br>(0.76 – 1.88) | 0.436 | | | | |
| Ertapenem | 0.41<br>(0.16 – 1.07) | 0.069 | 0.77<br>(0.51 – 1.17) | 0.222 | 0.36 $^{***}$<br>(0.23 – 0.58) | **<0.001** | | | | |
| Erythromycin | 12.87 $^{***}$<br>(5.75 – 28.81) | **<0.001** | 10.99 $^{***}$<br>(8.07 – 14.97) | **<0.001** | 25.22 $^{***}$<br>(18.68 – 34.06) | **<0.001** | | | | |
| Gentamicin | 4.36 $^{***}$<br>(2.50 – 7.57) | **<0.001** | 6.44 $^{***}$<br>(4.97 – 8.33) | **<0.001** | 3.14 $^{***}$<br>(2.41 – 4.08) | **<0.001** | | | | |
| Imipenem | 0.64<br>(0.33 – 1.23) | 0.182 | 0.72 $^{*}$<br>(0.54 – 0.96) | **0.028** | 0.36 $^{***}$<br>(0.25 – 0.53) | **<0.001** | | | | |
| Levofloxacin | 2.37 $^{**}$<br>(1.23 – 4.56) | **0.010** | 7.15 $^{***}$<br>(5.25 – 9.75) | **<0.001** | 6.69 $^{***}$<br>(4.98 – 8.99) | **<0.001** | | | | |
| Meropenem | 0.88<br>(0.38 – 2.02) | 0.757 | 0.87<br>(0.61 – 1.24) | 0.453 | 0.42 $^{***}$<br>(0.27 – 0.66) | **<0.001** | | | | |
| Penicillin.G | 65.33 $^{***}$<br>(19.20 – 222.24) | **<0.001** | 142.39 $^{***}$<br>(92.54 – 219.08) | **<0.001** | 22.37 $^{***}$<br>(16.24 – 30.83) | **<0.001** | | | | |
| Piperacillin | 15.32 $^{***}$<br>(8.74 – 26.85) | **<0.001** | 27.14 $^{***}$<br>(19.61 – 37.56) | **<0.001** | 25.76 $^{***}$<br>(19.41 – 34.18) | **<0.001** | | | | |
| Piperacillin.tazobactam | 5.01 $^{***}$<br>(2.50 – 10.02) | **<0.001** | 3.10 $^{***}$<br>(2.23 – 4.33) | **<0.001** | 2.55 $^{***}$<br>(1.77 – 3.68) | **<0.001** | | | | |
| Tetracycline | 4.10 $^{***}$<br>(2.18 – 7.71) | **<0.001** | 6.16 $^{***}$<br>(4.66 – 8.15) | **<0.001** | 6.76 $^{***}$<br>(5.12 – 8.93) | **<0.001** | | | | |
| TMPS | 10.94 $^{***}$<br>(6.36 – 18.82) | **<0.001** | 17.95 $^{***}$<br>(13.98 – 23.06) | **<0.001** | 24.28 $^{***}$<br>(18.86 – 31.27) | **<0.001** | | | | |

| | | | | | | | | | | |
|---|---|---|---|---|---|---|---|---|---|---|
| Vancomycin | 1.35 (0.44 – 4.13) | 0.595 | 2.65 *** (1.68 – 4.17) | **<0.001** | 4.34 *** (3.15 – 5.99) | **<0.001** | | | | |
| Q2 | 2.18 *** (1.65 – 2.90) | **<0.001** | 1.15 * (1.02 – 1.29) | **0.020** | 1.23 *** (1.11 – 1.36) | **<0.001** | | | | |
| Q3 | 1.72 *** (1.33 – 2.22) | **<0.001** | 1.29 *** (1.15 – 1.44) | **<0.001** | 1.17 ** (1.06 – 1.29) | **0.002** | | | | |
| Q4 | 1.78 *** (1.36 – 2.34) | **<0.001** | 1.50 *** (1.32 – 1.70) | **<0.001** | 1.19 ** (1.07 – 1.32) | **0.002** | | | | |
| Nitrofurantoin | | | | | 0.86 (0.63 – 1.17) | 0.343 | | | | |
| YEAR 2020 | | | | | | | 1.18 (0.90 – 1.55) | 0.227 | 1.27 (0.94 – 1.71) | 0.119 |
| YEAR 2021 | | | | | | | 1.22 (0.91 – 1.64) | 0.191 | 1.38 * (1.01 – 1.88) | **0.044** |
| Random Effects | | | | | | | | | | |
| $\sigma^2$ | 3.29 | | 3.29 | | 3.29 | | 3.29 | | 3.29 | |
| $\tau_{00}$ | 0.02 $_{YEAR}$ | | 0.05 $_{YEAR}$ | | 0.07 $_{YEAR}$ | | 0.06 $_{Classification}$ | | 0.09 $_{Classification}$ | |
| ICC | 0.01 | | 0.01 | | 0.02 | | 0.02 | | 0.03 | |
| N | 3 $_{YEAR}$ | | 3 $_{YEAR}$ | | 3 $_{YEAR}$ | | 2 $_{Classification}$ | | 2 $_{Classification}$ | |
| Observations | 2819 | | 13517 | | 16746 | | 2644 | | 2435 | |
| Marginal $R^2$ / Conditional $R^2$ | 0.351 / 0.355 | | 0.353 / 0.363 | | 0.341 / 0.354 | | 0.078 / 0.094 | | 0.150 / 0.172 | |

* *p<0.05* ** *p<0.01* *** *p<0.001*
